# A Binary Prototype for Time-Series Surveillance and Intervention

**DOI:** 10.1101/2025.02.03.25321613

**Authors:** Jason Olejarz, Till Hoffmann, Alex Zapf, Douaa Mugahid, Ross Molinaro, Chadwick Brown, Artem Boltyenkov, Taras Dudykevych, Ankit Gupta, Marc Lipsitch, Rifat Atun, Jukka-Pekka Onnela, Sarah Fortune, Rangarajan Sampath, Yonatan H. Grad

## Abstract

Despite much research on early detection of anomalies from surveillance data, a systematic framework for appropriately acting on these signals is lacking. We addressed this gap by formulating a hidden Markov-style model for time-series surveillance, where the system state, the observed data, and the decision rule are all binary. We incur a delayed cost, *c*, whenever the system is abnormal and no action is taken, or an immediate cost, *k*, with action, where *k* < *c*. If action costs are too high, then surveillance is detrimental, and intervention should never occur. If action costs are sufficiently low, then surveillance is detrimental, and intervention should always occur. Only when action costs are intermediate and surveillance costs are sufficiently low is surveillance beneficial. Our equations provide a framework for assessing which approach may apply under a range of scenarios and, if surveillance is warranted, facilitate methodical classification of intervention strategies. Our model thus offers a conceptual basis for designing real-world public health surveillance systems.

## Background

Temporal surveillance is integral to medicine and public health [1, 2]. Surveillance can be achieved using targeted sampling or through data that is already being collected for other purposes [3]. There are many potential data sources and diverse applications. Medical diagnostics devices are subject to quality control, and deviations from their designed behavior must be efficiently identified [4]. Hospitalized patients are constantly monitored in real time as part of their clinical care [5]. Healthy or chronically ill individuals are tested at the clinic at regular intervals, and decisions to change their treatment regimen must be deduced from this data [6].

In the context of pathogen control, syndromic surveillance considers the number of patients presenting with a constellation of symptoms. This data can be used to detect disease trends and outbreaks early and to inform if action is warranted [7, 8]. Active surveillance for pathogens includes wastewater sampling, which tracks the presence and abundance of a pathogen over time in the local sewershed, and monitoring for zoonotic diseases, where the vector population is regularly trapped and analyzed [9]. Public health organizations, such as the WHO and the CDC, are building large databases and are investing in substantial informatics infrastructure for intercepting infectious disease outbreaks [10]. Advancements in data science and machine learning have greatly expanded the potential for leveraging these systems to guide public health policy [11, 12].

For any scenario in which temporal surveillance is implemented, the purpose, therefore, is to use the signal to inform action [13, 14, 15]. If the time series indicates an abnormality in the sampled system that could lead to harm, then appropriate measures can be taken in response [16, 17, 18]. But how much deviation from some baseline is required to trigger an intervention [19, 20, 21]? If our threshold for acting is too high, then the condition or disease may spread before any countermeasures are taken, causing substantial harm. If our threshold is too low, then frequent overreactions result in accumulation of excessive intervention costs. The key point, then, is knowing when to intervene given a particular time-series signal.

## Model

We begin by identifying the basic components of the problem (Figure 1). The system state could correspond to the functioning of a medical device, the condition of a patient, or the presence of an infectious agent in a population. The system state is not directly known, but its effects are observed (perhaps at a later date) and contribute to the total cost, unless we take immediate action. Therefore, we construct a surveillance protocol, which also contributes to the total cost (even if only negligibly) and produces a data stream that provides real-time information on the system state. We further specify a decision rule for whether to intervene based on this data. Whenever we intervene, there is a contribution to the total cost.

**Figure 1.**
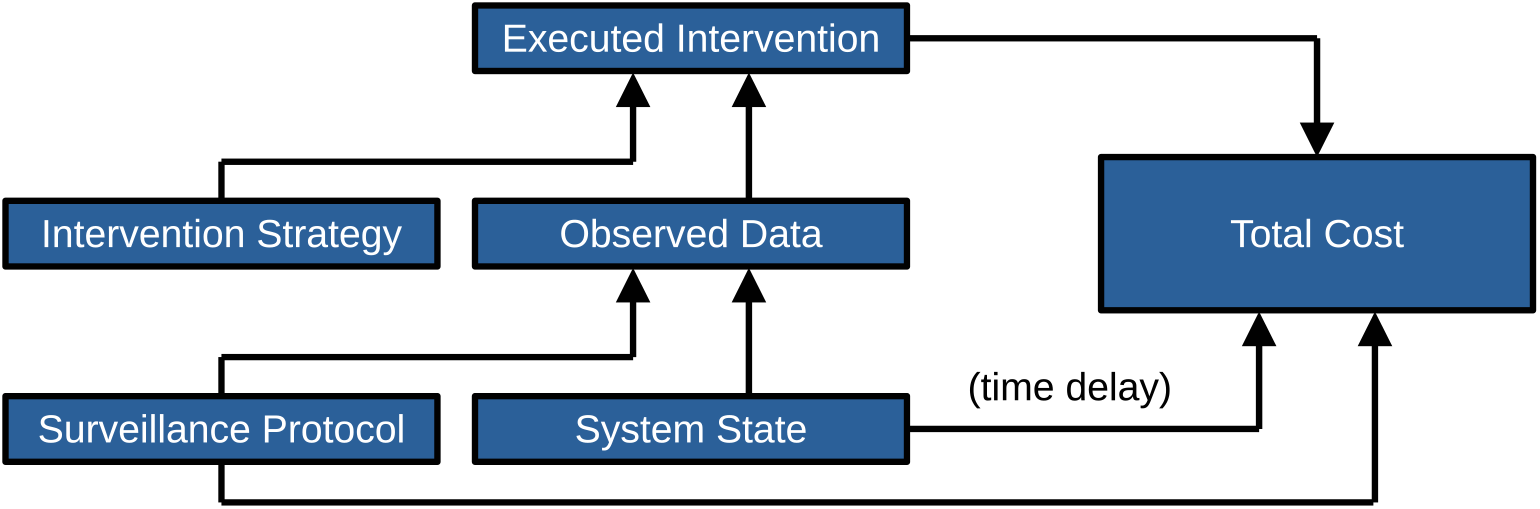
Building a surveillance and intervention platform. The system state affects both the total cost (after a time delay) and the observed data. The surveillance protocol affects both the total cost and the observed data. The observed data and the intervention strategy together determine whether an intervention is applied, and any executed intervention also contributes to the total cost.

The system state, the surveillance protocol, and the intervention strategy therefore determine the total cost, and we must choose the surveillance protocol and intervention strategy that minimize this quantity [22]. To make progress, we treat each of the three contributions to the total cost in its simplest manifestation. First, consider the system surveyed, which could be the operational integrity of a medical device, a patient’s biological or physiological condition, or the state of an entire population of individuals. The state of the system could typically be classified using many variables. In our model, however, we assume that the system at any given time is in one of two possible states—normal or abnormal—and we treat time as being discrete. The state of the system versus time is therefore a binary time series [23, 24]. If the system is in the abnormal state at a particular time, *t*, and no action is taken, then a resulting cost, *c*, is incurred. This could, for example, arise from operational failure of a diagnostics device, deterioration of a patient’s or population’s condition, or unmitigated spread of an infectious agent. If we take action at time *t*, then we instead incur a different cost, *k*. Notice that *k* encompasses both the costs due to the intervention and any remaining costs due to the abnormality despite having applied the intervention. (We assume that any costs incurred at one time step have no effect on costs incurred at the next. An example would be the costs incurred due to a vector-borne pathogen for which humans are dead-end hosts. In this case, spillover infections (and their associated costs) during one time interval are independent of spillover infections during the next. Similarly, if people are advised to stay indoors, then the associated intervention costs are felt only during the time interval in which they are employed.) Since the two possible responses at each time point are either action or inaction, the decisions to intervene or to not intervene also form a binary time series. The optimization problem presents when *k* < *c*. (Another way to state this is that *k* < *c* defines *c* to be a cost associated with an abnormality, since if that inequality does not hold, intervention in our model is ineffective.) We want to intervene if and only if the system is in the abnormal state, thereby averting harm from all threats without accumulating any unnecessary intervention costs (Figure 2).

**Figure 2.**
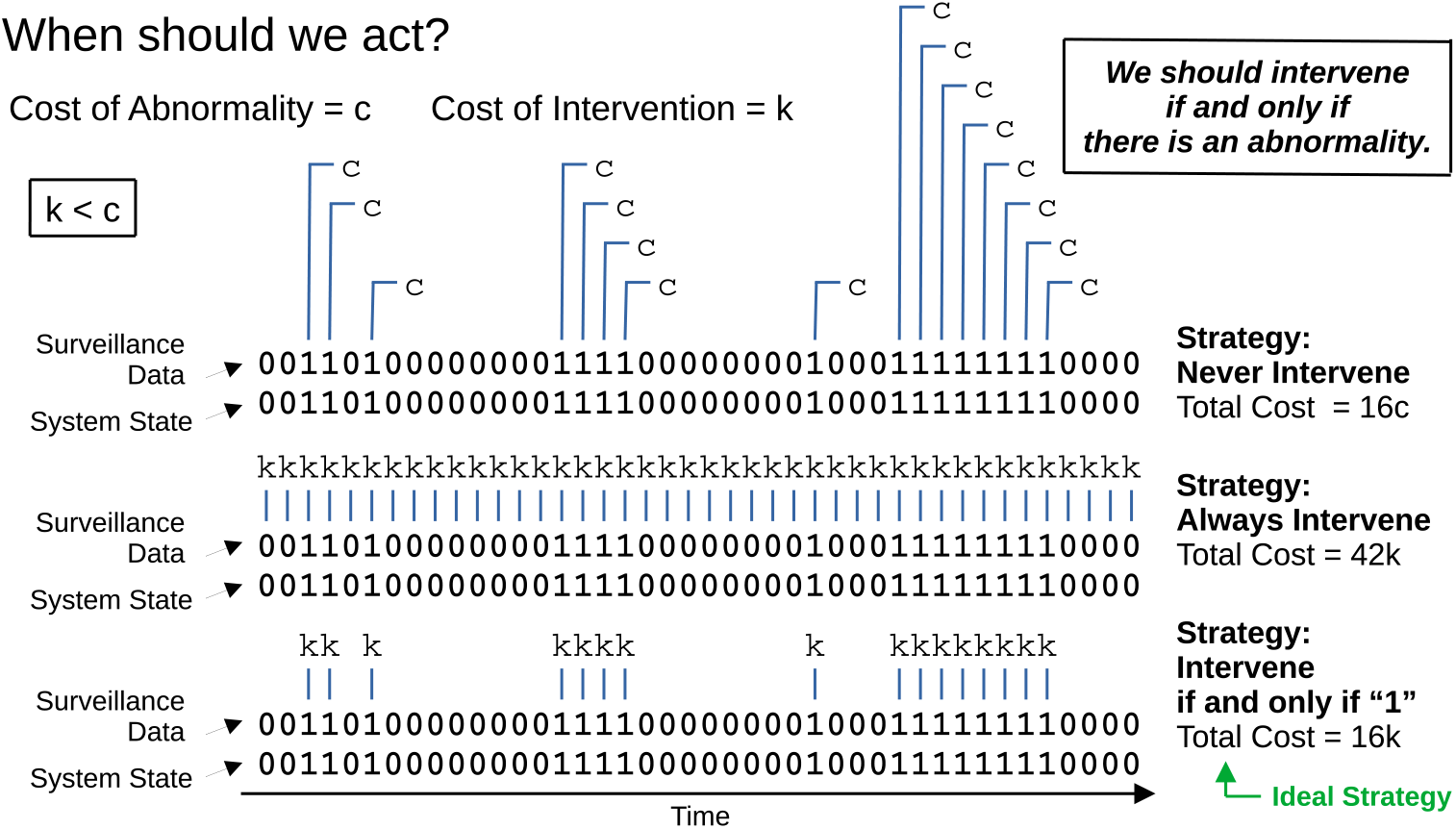
We would ideally intervene exactly when the system state is abnormal. At each time point, our system is in one of two states: normal or abnormal. If the system is normal, then there is no cost incurred related to any abnormality. If the system is abnormal and we don’t intervene, then a cost, *c*, is incurred. Whenever we intervene, we always incur a cost, *k*. For *k* < *c*, the optimal strategy is to intervene if and only if there is an abnormality.

To do that, we suppose that the observed data stream is a binary time series—i.e., at each time point, our surveillance program reads and reports a single bit of data, reflecting the system state as either normal or abnormal. The meaning of this bit can vary. If a device is being tested for proper functioning, then the bit reflects its state of operation. If a patient is being monitored, then the bit reflects the concentration of a particular biomarker. If a population is being sampled, then the bit reflects the number of clinical cases or abundance of an infectious agent in a defined geographic area. In all cases, there is a translation of a measured numeric value into a binary normal or abnormal indicator based on some threshold. Due to stochasticity, the imprecision of our measurement apparatus, and the imposition of our binary threshold, we neither directly nor accurately observe the state of the system, making our model similar to a hidden Markov model [25, 26].

Let *p*_00_ and *p*_01_ denote the probabilities of observing a 0 or 1, respectively, at time *t* given that the system is in the normal state at time *t*. Let *p*_10_ and *p*_11_ denote the probabilities of observing a 0 or 1, respectively, at time *t* given that the system is in the abnormal state at time *t*. Since our apparatus can only report a 0 or 1 at each measurement,

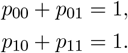

We assume these probabilities to be time-independent. Thus, the temporal data generated by our detector while the system is in a particular state is a Bernoulli sequence. We further consider that the abnormal state results in a higher probability of observing a 1 than the normal state:

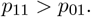

We postulate a simple rule for how the system can change. If the system is in the normal state at time *t*, then at time *t* + 1, it is in the abnormal state with probability *q*_01_ or the normal state with probability *q*_00_. If the system is in the abnormal state at time *t*, then at time *t* + 1, it is in the normal state with probability *q*_10_ or the abnormal state with probability *q*_11_. Since the state of the system at any given time is either normal or abnormal,

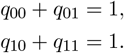

We assume these probabilities to be time-independent. Similarly, we assume that whenever we intervene, doing so does not alter the system state. (So, for example, we would not use this model for understanding when to impose a lockdown. We could, however, use this model for understanding when to recommend staying indoors to prevent infection with a vector-borne pathogen, particularly when humans are dead-end hosts. In this latter example, the intervention reduces the number of human infections, but the underlying dynamics of the pathogen in the vector population remain unperturbed.) Practically, for any particular application, these probabilities could be estimated from historical data or from independent modeling, which is beyond the scope of this work. We make one further assumption:

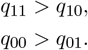

Thus, the system is more likely to remain in its present state at the next time point than to change states. With this constraint, the optimization of surveillance using two or more bits of data is a problem in reinforcement learning [27].

The task at hand is to use the resulting bit sequence to guide our intervention strategy. If *p*_00_ = 1 and *p*_11_ = 1, then we have exact knowledge of the state of the system, and the intervention strategy is to intervene if and only if we observe a 1. But such a scenario is an idealization. Realistically, we expect that *p*_00_ *<* 1 and *p*_11_ *<* 1—i.e., there are nonzero probabilities of both Type I and Type II errors (Figure S1). Whenever a Type I error occurs, we unnecessarily incur an intervention cost, *k*. Whenever a Type II error occurs, the delayed cost of inaction, *c*, exceeds the cost that we would have incurred had we intervened, *k*. We therefore want to minimize Type I and Type II errors (Figure S2). How should we decide what to do in response to a noisy temporal signal?

**Figure S1:**
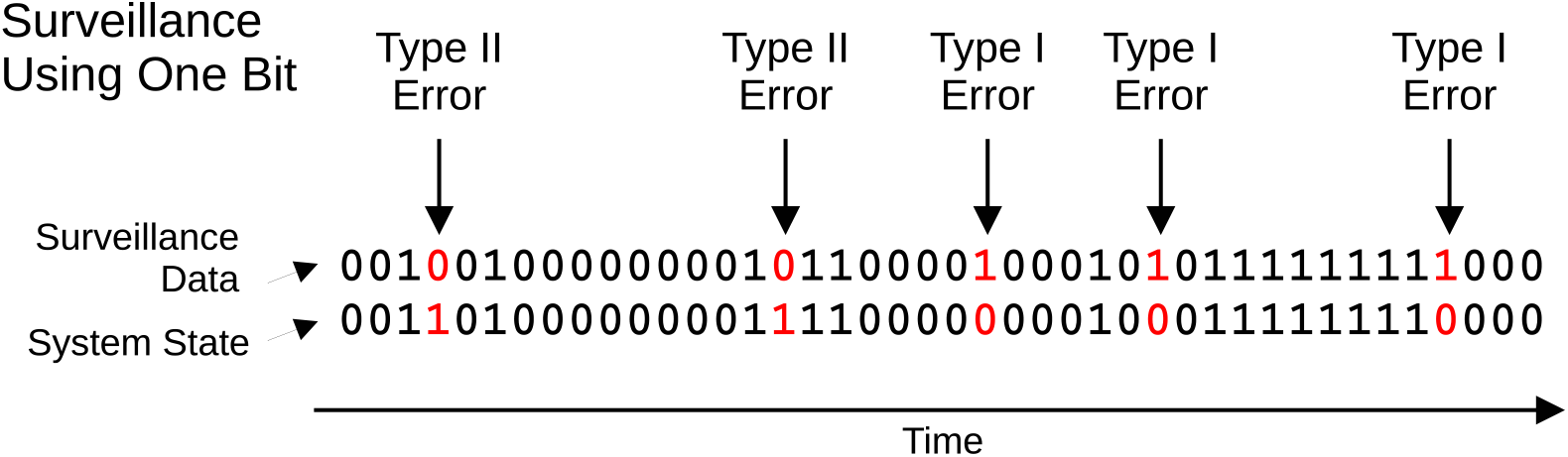
Measurement errors. Suppose that our intervention strategy is based on using only the most recent bit of data. A Type I error occurs whenever the system state is 0 and our diagnostics machine reports 1. A Type II error occurs whenever the system state is 1 and our diagnostics machine reports 0.

**Figure S2:**
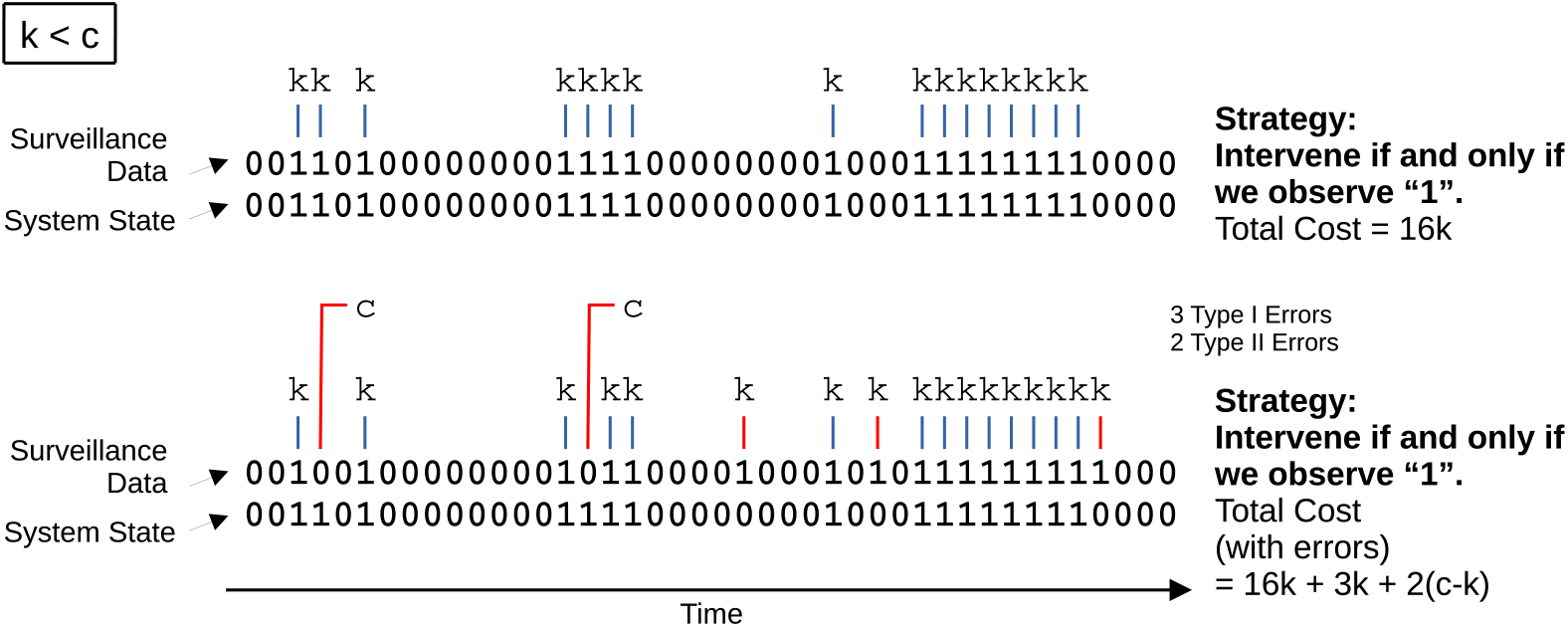
Accounting for measurement errors in the calculation of total cost. Whenever a Type I error occurs, we unnecessarily intervene and incur a cost, *k*. Whenever a Type II error occurs, we incur a cost, *c*, at a later time instead of having incurred a smaller, immediate cost *k* had we intervened.

For obtaining concrete results and building intuition, we make one further simplification. Consider *a*_1_(*t*), which is the probability that the system is in the abnormal state at a particular time, *t*. If the system is in the abnormal state at time *t*, then one of two things happened: Either the system was already in the abnormal state at time *t* − 1 and didn’t transition to the normal state, or the system was in the normal state at time *t* − 1 and transitioned to the abnormal state. This leads to the following recurrence relation:

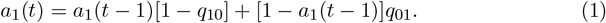

We simplify this by considering that the system is in steady state—i.e., that the stochastic dynamics have run for sufficiently long that the probability that the system is in the abnormal state is time-independent. Substituting *a*_1_(*t*) → *a*_1_ into Equation (1) and solving for *a*_1_, we obtain

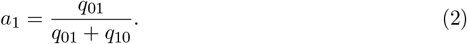

Therefore, our prior—i.e., the probability that the system is in the abnormal state at any given time in the absence of any other information—is known exactly and is specified by Equation (2). We also define *a*_0_ to be the probability that the system is in the normal state at any given time, so that

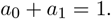

Bayesian inference thus yields the exact probability that the system is in the abnormal state given the most recent *n* bits of data [28, 29]. By formally specifying a loss function that accounts for all possible types of costs and then minimizing this function, we can determine the ideal surveillance and intervention strategy.

All parameters are shown in Table 1.

**Table 1:** Table of parameters.

| Basic Parameters |  |
| --- | --- |
| $p_{00}$ | Probability of observing 0 while in the normal state |
| $p_{01}$ | Probability of observing 1 while in the normal state |
| $p_{10}$ | Probability of observing 0 while in the abnormal state |
| $p_{11}$ | Probability of observing 1 while in the abnormal state |
| $q_{00}$ | Probability of remaining in the normal state |
| $q_{01}$ | Probability of switching from the normal state to the abnormal state |
| $q_{11}$ | Probability of remaining in the abnormal state |
| $q_{10}$ | Probability of switching from the abnormal state to the normal state |
| $c$ | Cost due to an abnormality that is not acted on |
| $k$ | Intervention cost |
| $s_n$ | Surveillance cost per unit time using $n$ bits of data |
| Derived Parameters |  |
| $a_0 = q_{10} / (q_{10} + q_{01})$ | Exact prior probability of being in the normal state |
| $a_1 = q_{01} / (q_{10} + q_{01})$ | Exact prior probability of being in the abnormal state |
| $K = k / c$ | Normalized intervention cost |
| $S_n = s_n / c$ | Normalized surveillance cost per unit time using $n$ bits of data |

## Applications

To demonstrate the protocol, we first suppose that there is no surveillance. This serves as a baseline from which to gauge the effects of different surveillance and intervention strategies. We then explore how using a single bit of data from our detector to guide our intervention can be beneficial. We next explore the use of two bits of data from our detector, and we outline a methodology for evaluating progressively more complex surveillance and intervention protocols. For all calculations, we use the steady-state assumption of Equation (2). We also consider all costs to be measured relative to the cost of an abnormality that is not acted on.

### No surveillance

Consider first the simplest possibility: that there is no surveillance at all. Let *K* = *k/c*. If the system is typically in the normal state, or if *K* is sufficiently high, then intervening is too costly. But if there are frequent transitions to the abnormal state, or if *K* is sufficiently low, then intervening might be cost-effective. How should we decide what to do? If we always intervene, then the cost incurred per unit time, which we denote *L*_0_(1), is equal to *K*. If we never intervene, then the expected cost incurred per unit time, which we denote *L*_0_(0), is equal to *a*_1_. Therefore, selecting between these two options, the optimal strategy is to always intervene if *a*_1_ *> K* and never intervene otherwise. The intervention decision is thus solely informed by cost considerations.

### Surveillance using one bit of data

We extend this framework to the next-simplest case: that we use the single most recent bit of data from our surveillance apparatus to inform our intervention strategy. Let *Y*_0_ and *Y*_1_ represent the intervention strategy given that the most recent bit is 0 or 1, respectively. If *Y*_0_ = 1, then we always intervene when we observe a 0, and if *Y*_0_ = 0, then we never intervene when we observe a 0. Likewise, if *Y*_1_ = 1, then we always intervene when we observe a 1, and if *Y*_1_ = 0, then we never intervene when we observe a 1. Let *S*_1_ = *s*_1_*/c* denote the cost per unit time of implementing surveillance using one bit of data. The expected cost per unit time is equal to

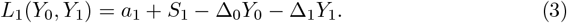

Here, as shorthand notation, we have defined

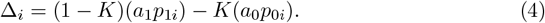

(Δ_*i*_ is the expected cost per unit time of not intervening when we observe *i* minus the expected cost per unit time of intervening when we observe *i*.) The ideal intervention strategy when using one bit of data is the one for which *L*_1_(*Y*_0_, *Y*_1_) is minimized. The consideration is whether Δ_0_ and Δ_1_, given by Equations (4), are positive or negative. From Equation (3), if Δ_0_ > 0 or Δ_0_ < 0, then we should set *Y*_0_ = 1 or *Y*_0_ = 0, respectively. Likewise, if Δ_1_ > 0 or Δ_1_ < 0, then we should set *Y*_1_ = 1 or *Y*_1_ = 0, respectively. The optimal intervention strategy is then

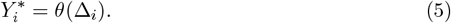

Here, *θ* denotes the Heaviside step function. Substituting 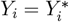, given by Equations (5), into Equations (3), we minimize *L*_1_(*Y*_0_, *Y*_1_).

Considering that we can use either no surveillance or surveillance based on the most recent bit, there are six possible strategies and six corresponding loss functions: *L*_0_(0), *L*_0_(1), *L*_1_(0, 0), *L*_1_(1, 0), *L*_1_(0, 1), and *L*_1_(1, 1). But the analysis simplifies. *L*_1_(0, 1) < *L*_1_(1, 0) is necessarily true, since an observation of 1 in the most recent bit indicates a higher probability of being in the abnormal state. Also, *L*_0_(*Y*) < *L*_1_(*Y, Y*) is necessarily true, since using the most recent bit of data entails a surveillance cost per unit time of *S*_1_, whereas using no data entails no surveillance cost. There are then three possibilities: We can choose to (i) never intervene, (ii) always intervene, or (iii) intervene if and only if the most recent bit is 1. We should choose the first option if *L*_0_(0) < *L*_0_(1) and *L*_0_(0) < *L*_1_(0, 1). We should choose the second option if *L*_0_(1) < *L*_0_(0) and *L*_0_(1) < *L*_1_(0, 1). We should choose the third option if *L*_1_(0, 1) < *L*_0_(0) and *L*_1_(0, 1) < *L*_0_(1). The numerical optimization problem using one bit of data is illustrated in Table S1. Optimization of surveillance using one bit of data is summarized in Figure 3. An example that involves optimizing surveillance for vector-borne pathogens is given in Figure S3.

**Figure 3.**
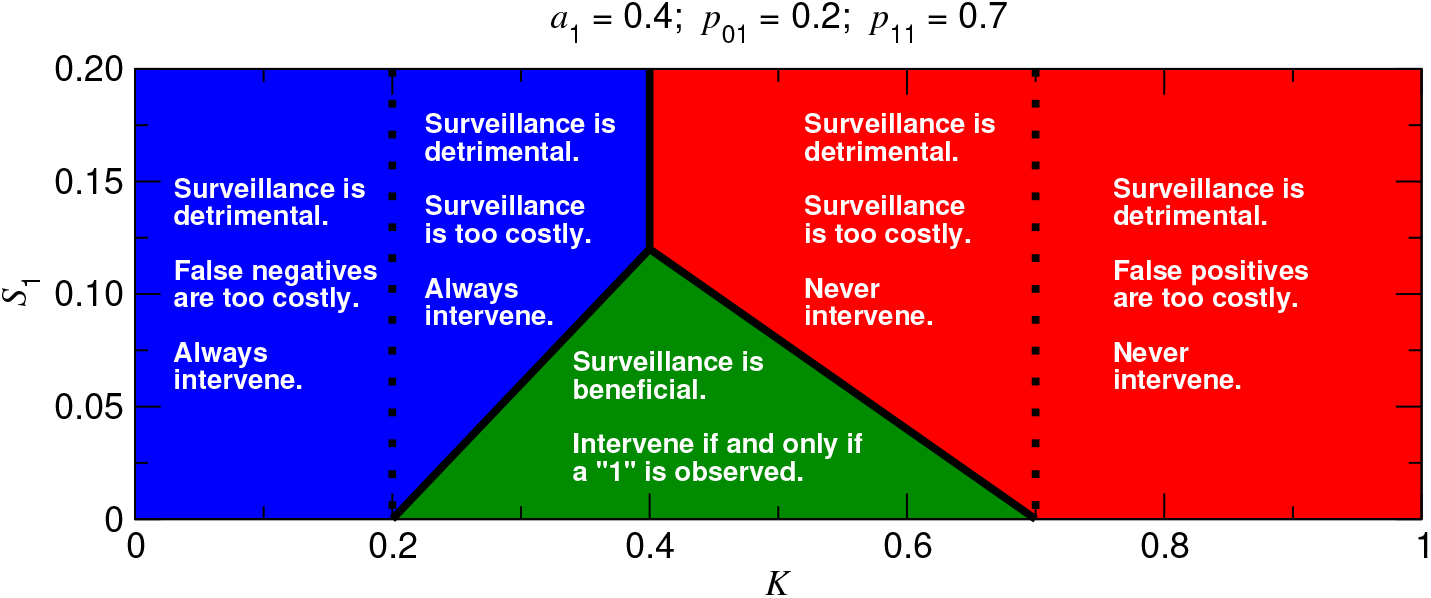
Surveillance using one bit of data. The decision of whether to use surveillance based on the most recent bit of data depends on the values of *K* and *S*_1_. In the red region, we should not use surveillance, and we should never intervene. In the blue region, we should not use surveillance, and we should always intervene as standard practice. Surveillance should only be implemented within the green triangle. (*a*_1_: exact prior probability of being in the abnormal state; *p*_01_: probability of observing 1 while normal; *p*_11_: probability of observing 1 while abnormal.)

### Surveillance using two bits of data

Extending this framework further, suppose that we use the two most recent bits of data from our surveillance apparatus to inform our intervention strategy. Let *Y*_*ij*_ represent the intervention strategy given that the most recent bit is *j* and the bit before that is *i*. If *Y*_*ij*_ = 1, then we always intervene when we observe *ij*, and if *Y*_*ij*_ = 0, then we never intervene when we observe *ij*. Let *S*_2_ = *s*_2_*/c* denote the cost per unit time of implementing surveillance using two bits of data. The expected cost per unit time is equal to

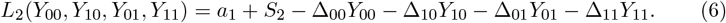

Here, as shorthand notation, we have defined

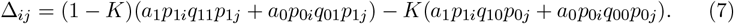

(Δ_*ij*_ is the expected cost per unit time of not intervening when we observe *ij* minus the expected cost per unit time of intervening when we observe *ij*.) The ideal intervention strategy when using two bits of data is the one for which *L*_2_(*Y*_00_, *Y*_10_, *Y*_01_, *Y*_11_) is minimized. The consideration is whether Δ_*ij*_, given by Equations (7), are positive or negative. From Equations (6), if Δ_*ij*_ > 0 or Δ_*ij*_ < 0, then we should set *Y*_*ij*_ = 1 or *Y*_*ij*_ = 0, respectively. The optimal intervention strategy is then

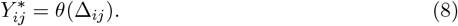

Substituting 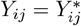, given by Equations (8), into Equations (6), we minimize *L*_2_(*Y*_00_, *Y*_10_, *Y*_01_, *Y*_11_).

When the possibility of using two bits of data is considered, optimization of surveillance becomes a more involved calculation (Figures S4 and S5). To understand how to optimize surveillance using two bits of data, we must consider the values of *K, S*_1_, and *S*_2_. There are five possibilities: We can choose to (i) never intervene, (ii) always intervene, (iii) intervene if and only if the most recent bit is 1, (iv) intervene if and only if the two most recent bits are 1, or (v) intervene if and only if at least one of the two most recent bits is 1. For determining which surveillance and intervention strategy is best, there are thus five loss functions to consider: *L*_0_(0), *L*_0_(1), *L*_1_(0, 1), *L*_2_(0, 0, 0, 1), and *L*_2_(0, 1, 1, 1). We should choose the first strategy if *L*_0_(0) is lowest, the second strategy if *L*_0_(1) is lowest, the third strategy if *L*_1_(0, 1) is lowest, the fourth strategy if *L*_2_(0, 0, 0, 1) is lowest, or the fifth strategy if *L*_2_(0, 1, 1, 1) is lowest. The numerical optimization problem using two bits of data is illustrated in Table S2. Optimization of surveillance using two bits of data is summarized in Figure 4. An example of using two bits of data for optimizing surveillance for vector-borne pathogens is given in Figure S6.

**Figure 4.**
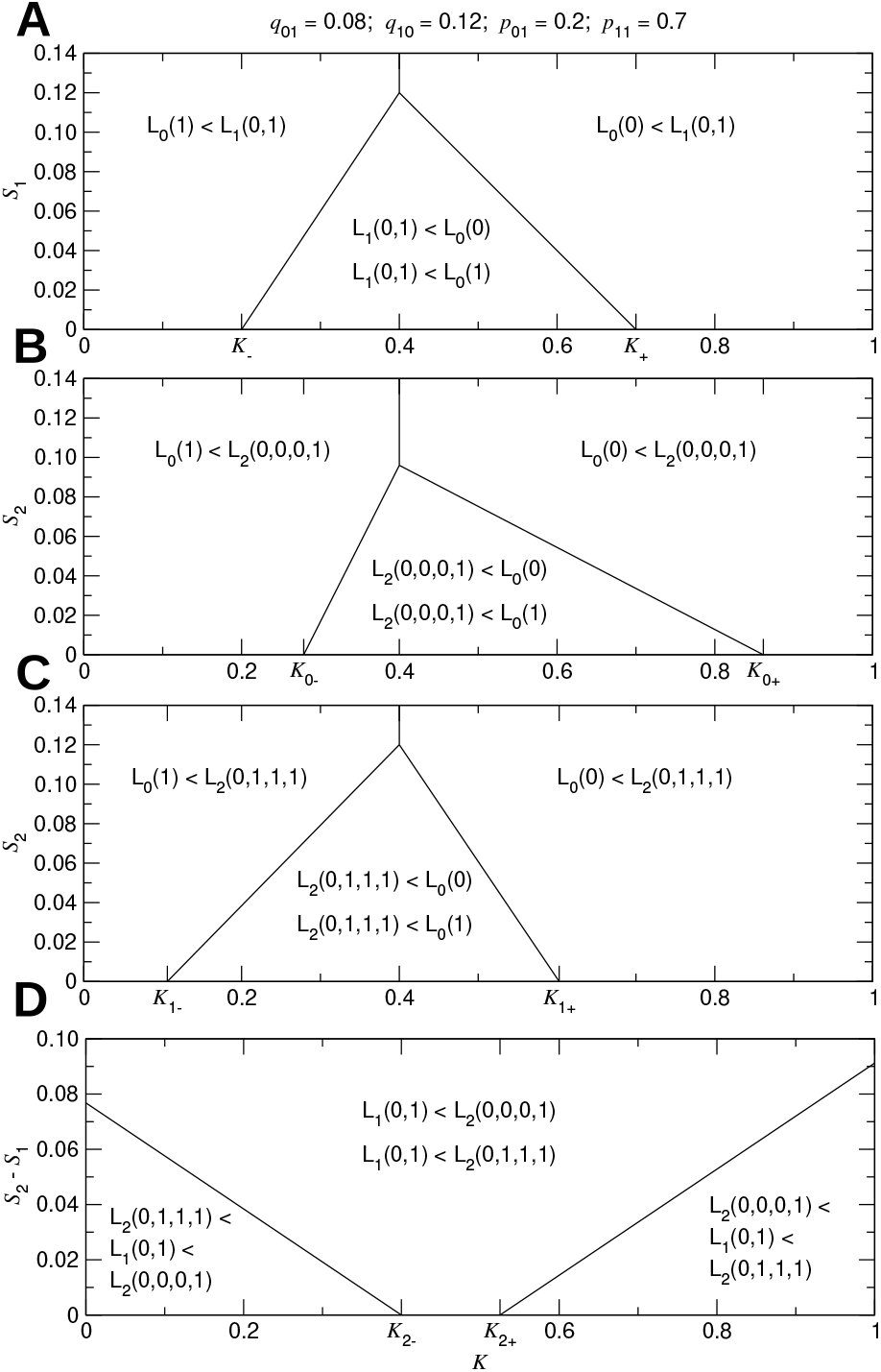
Surveillance using two bits of data. For this example, we consider five possibilities: (i) no surveillance; never intervene, (ii) no surveillance; always intervene, (iii) surveillance using one bit; intervene if and only if the most recent bit is 1, (iv) surveillance using two bits; intervene if and only if both of the two most recent bits are 1, or (v) surveillance using two bits; intervene if and only if at least one of the two most recent bits is 1. We compare strategies (i), (ii), and (iii) in (A), strategies (i), (ii), and (iv) in (B), strategies (i), (ii), and (v) in (C), and strategies (iii), (iv), and (v) in (D). The ideal surveillance and intervention strategy is the one for which the corresponding cost function among the five considered possibilities is lowest, which is determined by the values of *K, S*_1_, and *S*_2_. (*K*: normalized intervention cost; *S*_*n*_: normalized surveillance cost per unit time using *n* bits of data; *q*_01_: probability of switching from normal to abnormal; *q*_10_: probability of switching from abnormal to normal; *p*_01_: probability of observing 1 while normal; *p*_11_: probability of observing 1 while abnormal.)

### Surveillance using any number of bits of data

The steps in the calculations are the same if we use any number of bits, *n*, for surveillance. Let 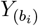 represent the intervention strategy given that the most recent sequence of bits is 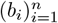. If 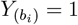, then we always intervene when we observe 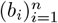, and if 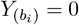, then we never intervene when we observe 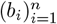. Let *S*_*n*_ = *s*_*n*_*/c* denote the cost per unit time of implementing surveillance using *n* bits of data. For *n* ≥ 1, the expected cost per unit time is equal to

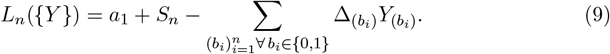

Here, as shorthand notation, we have defined

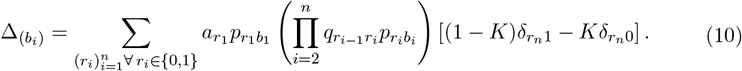

In Equations (10), *δ*_*ij*_ denotes the Kronecker delta. The ideal intervention strategy when using *n* bits of data is the one for which *L*_*n*_({*Y*}) is minimized. The consideration is whether each 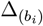, given by Equations (10), is positive or negative. From Equations (9), if 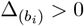 or 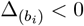, then we should set 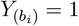 or 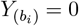, respectively. If we use 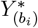 to represent the optimal intervention strategy given that the *n* most recent bits are 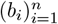, then we have

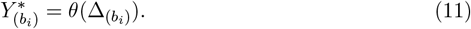

Substituting 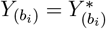, given by Equations (11), into Equations (9), we minimize *L*_*n*_({*Y*}).

There are many possible scenarios to consider. For example, suppose that we are using the three most recent bits of data to inform our intervention strategy. If the three most recent bits are 001, does the most recent observation of 1 outweigh the two prior observations of 00, or vice versa? Similarly, if the three most recent bits are 110, does the most recent observation of 0 outweigh the two prior observations of 11, or vice versa? Figure S7 shows that the answers to these questions are subtle and dependent on parameter values. We might also ask if using three bits of data in the decision to intervene is justified. For certain parameter values, surveillance using three bits of data delivers a large reduction in the expected cost per unit time versus using only two bits of data (Figure S8). Optimizing surveillance using more than two bits of data must therefore be done carefully.

## Discussion

Hidden Markov models have been studied in diverse contexts and for many applications. They are valuable tools for the optimization of quality control, such as in fault detection [30, 31, 32, 33], degradation assessment [34, 35, 36], predictive maintenance [37, 38], determination of water quality [39, 40], and pharmaceutical process monitoring [41]. They have important uses in medical diagnostics surveillance, including analyzing changes in gene expression [42], testing for drug effectiveness [43], checking cardiovascular function [44], diagnosing respiratory illness [45], managing diabetes [46], and screening for cancer [47]. They have also been proposed for interpreting public health surveillance data [48], with demonstrated uses in understanding nosocomial pathogen transmission [49], tuberculosis cases [50], influenza epidemics [51, 52, 53], seasonal epidemics [54, 55], COVID-19 data [56], and other types of infectious disease outbreaks [57, 58, 59, 60, 61]. Inspired by these applications, our work fills a critical void by assigning a cost to an abnormality, a cost per unit time to the surveillance protocol, and a cost to any executed intervention. This advancement precisely frames the optimization problem: Which surveillance protocol and intervention strategy results in the lowest expected cost per unit time?

Although our model is simple, it is broadly adaptable. Our assumption that the system state is binary is a valid first step in many settings. For example, a piece of equipment might normally operate around a stable equilibrium point (system state 0) and only begin to malfunction after a large perturbation forces it toward another stable equilibrium point (system state 1). A patient might be healthy for a long period (system state 0) before their condition suddenly worsens (system state 1). Or an infectious agent might be absent from a population for a long period (system state 0) and then spread rapidly following its introduction (system state 1). Our assumption that each measurement is binary could reflect a diagnostics device that only outputs two possible values, or it could result from dichotomizing a count variable using a suitable smoothing or preprocessing routine [62]. It could also reflect an oversimplification dictated by the realities of using data from a highly regulated industry. Our assumption that the intervention strategy is binary reflects the simplest possible type of intervention and is a reasonable starting point for any such analysis. The interpretation of time in the model can be adapted to the particular application at hand. For example, for reporting the prevalence of an infectious disease, each observation might reflect the number of new cases in a given week. The cost parameters are also adaptable and can include a wide variety of effects.

For this type of mathematical optimization to be effective, a suitable cost function to be minimized must be constructed. This is challenging, particularly since the system state and data reporting are ever-changing, complex processes that would not be reliably described by a small set of transition rules and parameters. A realistic implementation of this approach would be adaptive, requiring both inference of the rules governing the underlying process and prediction of an abnormality using the same data stream [63, 64, 65]. A further challenge is that the most harmful threats appear stochastically and at irregular intervals. This could be handled by including an additional cost parameter that is necessarily larger when using an intervention strategy that is less aggressive. This generates an incentive to be preemptive in reacting to abnormalities in the data—i.e., we tolerate higher intervention costs in exchange for the lessened probability of being devastated by a disturbance that grows uncontrollably.

Complicating this further is the variety of types of costs at play. If quality control is being performed on a device or manufacturing process, then intervention might entail fixing components or altering specifications that are provided to customers. If a patient is tested at the clinic, then intervention might mean changing the patient’s prescribed medications or suggesting modifications to their lifestyle. If an infectious disease is spreading, then intervention might involve imposing quarantines, encouraging masking, or developing vaccines. Although understanding these myriad costs using the same cost unit (e.g., time, money, effort, mental well-being, physical well-being, lives lost, etc.) seems daunting, determination of the optimal surveillance and intervention strategy requires it, since the parameters *S*_*n*_ and *K* must be dimensionless. The necessity of a unified understanding of costs persists as more complex and realistic models of this sort are investigated.

Even with the uncertainties inherent in assignment of costs, our model is critical for guiding public health policy. Regardless of rationale or justification, a practitioner necessarily believes that the surveillance and intervention protocol that they are using is optimal. When conditions change—e.g., if more people are presenting to the hospital, or if an arbovirus’s prevalence in the environment is increasing—a corresponding alteration to the surveillance and intervention program is warranted. Our model accordingly enables relative comparisons between strategies given different assumptions, and definitive recommendations to the public health practitioner on adaptations that should be made can be offered. In a similar vein, surveillance and intervention strategies differ geographically, again based on different location-specific observations and assumptions. If a particular policy is believed to be optimal for one location, then, based on different observations and assumptions, our model suggests suitable changes that should be made for another location.

Our methodology can be further developed and applied to complex real-world scenarios, where the state of the system is not binary. For example, equipment malfunctions can progress through stages such as warnings or partial functionality before full failure. Similarly, a patient’s health condition could range from mild to moderate to severe, and disease spread often involves multiple stages, such as latent, early spread, exponential, or stabilization. In a natural extension of this framework, one might accordingly consider that after a disturbance is initiated, its magnitude grows in time according to a branching process [66, 67]. If we wait longer before responding to the abnormality, then we incur a larger cost due to its effects. One might further suppose that our detector outputs a count at each time point, and we must use this data to inform our belief of whether a disturbance is present and whether to intervene. The optimization problem works the same: If our threshold for acting is too low, then the small expected size of an abnormality when we intervene does not justify the frequent intervention costs that are incurred. If our threshold for acting is too high, then an abnormality would typically grow to a large size before intervention is applied, and the intervention would only have minimal effectiveness. There might also be multiple disturbances of various magnitudes overlapping with each other, which could increase the chances of Type I and Type II errors. The task is therefore to determine whether we should intervene given a particular sequence of recent count measurements. This is an important direction for future work.

We have constructed a simple model for time-series surveillance and investigated its implications. A salient point is that surveillance can be either beneficial or detrimental, depending on many factors. Proposed applications of surveillance must be properly vetted for their usefulness before substantial resources are allocated to their development. Once a surveillance system is built, it is not enough to be able to identify or predict anomalies: Any observed signal must be integrated within a cost-benefit framework for understanding the consequences of particular responses to surveillance data. The surveillance system can only be meaningfully optimized by constructing a suitable cost function—with all cost parameters interpreted using a common cost unit—and identifying the intervention strategies for which this function is minimal. Even if precise assignment of costs is difficult, our model facilitates critical comparisons between strategies based on differing assumptions. Our model is a robust prototype for the effective implementation of surveillance programs that are used to inform public health policy.

## Data Availability

There are no data.

## Acknowledgements

JO is grateful for discussions with David Helekal. All authors were funded in whole or in part by Siemens Healthcare Diagnostics, Inc. (Siemens Healthineers-Harvard Chan RCA, 8317147-01). This project has been funded in part by contract 200-2016-91779 with the Centers for Disease Control and Prevention, United States (CDC). Disclaimer: The findings, conclusions, and views expressed are those of the author(s) and do not necessarily represent the official position of the CDC. ML is supported by cooperative agreement U01 CA261277 through the National Cancer Institute/US National Institutes of Health.

## Supplementary Information

This Supplementary Information is organized as follows. In Section 1, we derive the optimal intervention strategy given that there is no surveillance. In Section 2, we derive the optimal surveillance and intervention strategy using the most recent bit of data. We also give an example, where we apply our framework to optimize surveillance for vector-borne pathogens. In Section 3, we derive the optimal surveillance and intervention strategy using the two most recent bits of data. We also expand on the example of the previous section. In Section 4, we explore the optimization of surveillance and intervention using the three most recent bits of data. We further demonstrate some of the intricacies that arise when optimizing surveillance using three or more bits of data.

### 1 No surveillance

If there is no surveillance, then the optimal strategy is either to never intervene or to always intervene. If we never intervene, then we incur an expected cost per unit time equal to *a*_1_*c*. If we always intervene, then we incur a cost per unit time equal to *k*. Let *Y* represent the intervention strategy, so that *Y* = 0 if we never intervene and *Y* = 1 if we always intervene. The unnormalized expected cost per unit time,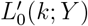, can be expressed as

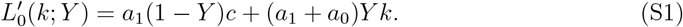

Letting *K* = *k/c* and 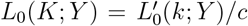 in Equation (S1), we obtain the normalized expected cost per unit time:

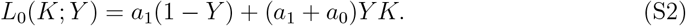

Using *a*_1_ + *a*_0_ = 1 in Equation (S2) and simplifying, we obtain

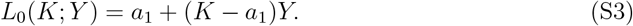

Equation (S3) serves as a baseline for gauging the effectiveness of more complex surveillance and intervention strategies.

### 2 Surveillance using one bit of data

The unnormalized expected cost per unit time given that we use the single most recent bit of data to inform our intervention strategy, 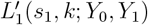, can be expressed as

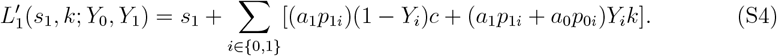

The terms in the summation in Equation (S4) have a simple interpretation. If we do not intervene at time *t* and the system is in the abnormal state at time *t*, then we incur a delayed cost, *c*. Considering the most recent bit of data, there are two ways that this can happen:

- The system can be in the abnormal state at time *t*, be measured as bit *i* at time *t*, and not be acted on at time *t*. This sequence of events occurs with probability *a*_1_*p*_1*i*_(1 −*Y*_*i*_), and *i* can be either 0 or 1.

If we intervene at time *t*, then we incur an immediate cost, *k*. Considering the most recent bit of data, there are four ways that this can happen:

- The system can be in the abnormal state at time *t*, be measured as bit *i* at time *t*, and be acted on at time *t*. This sequence of events occurs with probability *a*_1_*p*_1*i*_*Y*_*i*_, and *i* can be either 0 or 1.
- The system can be in the normal state at time *t*, be measured as bit *i* at time *t*, and be acted on at time *t*. This sequence of events occurs with probability *a*_0_*p*_0*i*_*Y*_*i*_, and *i* can be either 0 or 1.

Letting *K* = *k/c, S*_1_ = *s*_1_*/c*, and 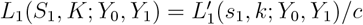 in Equation (S4), we obtain the normalized expected cost per unit time:

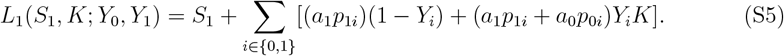

As shorthand notation, we define

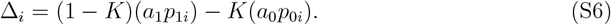

Rearranging Equation (S5), and using Equation (S6), we obtain

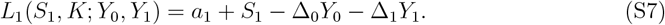

The task at hand is to use Equation (S7) and the observed bit sequence to guide our intervention strategy.

If *p*_01_ = 0 and *p*_11_ = 1, then we have exact knowledge of the state of the system, and the ideal intervention strategy is to intervene if and only if we observe a 1. But such a scenario is an idealization. Realistically, we expect that *p*_01_ > 0 and *p*_11_ < 1—i.e., there are nonzero probabilities of both Type I and Type II errors (Figure S1). Whenever a Type I error occurs, we unnecessarily incur an intervention cost, *k*. Whenever a Type II error occurs, the delayed cost of inaction, *c*, exceeds the immediate cost that we would have incurred had we intervened, *k*. We therefore want to minimize Type I and Type II errors (Figure S2). How should we decide what to do in response to a noisy temporal signal?

It is helpful to understand the ideal surveillance and intervention strategy depending on the values of *K* and *S*_1_ (Figure 3). If *K* is larger than a certain value, which we denote *K*_+_, then we should never intervene. *K*_+_ is given by solving *L*_1_(0, *K*_+_; 0, 1) = *L*_0_(*K*_+_; 0). We find

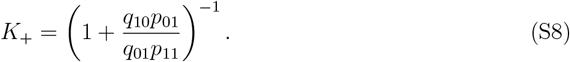

If *K* > *K*_+_, then the expected cost from reacting to false positives exceeds any benefit from averting the threat. The conclusion is that surveillance should not be executed, and intervention should never occur. *K*_+_ is shown as the right dotted vertical line in Figure 3.

Similarly, if *K* is smaller than a certain value, which we denote *K*_−_, then we should always intervene. *K*_−_ is given by solving *L*_1_(0, *K*_−_; 0, 1) = *L*_0_(*K*_−_; 1). We find

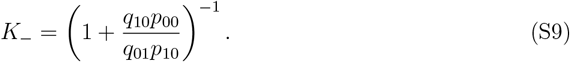

If *K* < *K*_−_, then the expected cost from dismissing false negatives exceeds any benefit from not intervening when there is no threat. The conclusion is that surveillance should not be executed, and intervention should always occur. *K*_−_ is shown as the left dotted vertical line in Figure 3.

*K*_+_ and *K*_−_, given by Equations (S8) and (S9), respectively, are therefore key quantities for determining if surveillance is beneficial. If *K*_−_ < *K* < *K*_+_, then surveillance might be justified if surveillance costs are sufficiently low. If we use surveillance, then we incur an expected cost per unit time equal to *L*_1_(*S*_1_, *K*; 0, 1). First, consider that *K* > *a*_1_. If we don’t use surveillance, then the optimal strategy is to never intervene, and we incur an expected cost per unit time equal to *L*_0_(*K*; 0). We solve for the value of *S*_1_ = *S*_+_ for which *L*_0_(*K*; 0) = *L*_1_(*S*_+_, *K*; 0, 1):

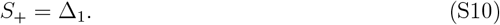

If *S*_1_ < *S*_+_, then the benefit of appropriately intervening outweighs the cost of surveillance, and surveillance is beneficial. If *S*_1_ > *S*_+_, then surveillance is too expensive and is detrimental. From Equation (S10), *S*_+_ is plotted versus *K* as the upper right boundary of the green triangular region in Figure 3.

Next, consider that *K* < *a*_1_. If we don’t use surveillance, then the optimal strategy is to always intervene, and we incur an expected cost per unit time equal to *L*_0_(*K*; 1). We solve for the value of *S*_1_ = *S*_−_ for which *L*_0_(*K*; 1) = *L*_1_(*S*_−_, *K*; 0, 1):

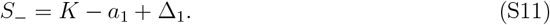

If *S*_1_ < *S*_−_, then the benefit of appropriately intervening outweighs the cost of surveillance, and surveillance is beneficial. If *S*_1_ > *S*_−_, then surveillance is too expensive and is detrimental. From Equation (S11), *S*_−_ is plotted versus *K* as the upper left boundary of the green triangular region in Figure 3.

**Table 1:**
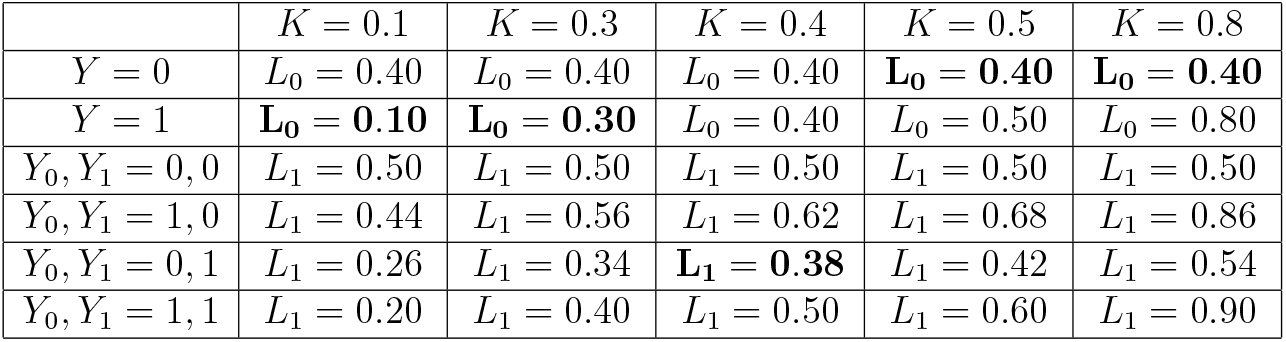
Example of optimizing surveillance and intervention using the most recent bit of data. We set *a*_1_ = 0.4, *p*_01_ = 0.2, *p*_11_ = 0.7, and *S*_1_ = 0.1.

#### 2.1 Example using one bit of data

A simple example helps for putting the model and its potential applications into context. Many vector-borne pathogens, such as West Nile virus (WNV) and eastern equine encephalitis virus (EEEV), persist in the environment and are characterized by sudden, intermittent outbreaks. The timing of these outbreaks is extremely difficult to predict. To model this, suppose that *q*_01_—defined for this example as the probability of an outbreak beginning in any given week—is small (*q*_01_ ≪ 1). It is also realistic to suppose that outbreaks do not suddenly and spontaneously go away (*q*_10_ ≪ 1).

Suppose that the vector population is sampled and tested weekly, and we measure the abundance of viral RNA, *v*, in each vector sample. When there is no outbreak, the abundance of viral RNA in each measurement is drawn from a probability distribution, *F*_0_(*v*). For each measurement during an ongoing outbreak, the abundance of viral RNA is drawn from a different probability distribution, *F*_1_(*v*). We define a cutoff value for the measured abundance of viral RNA in a sample, *v*′. The distributions *F*_0_(*v*) and *F*_1_(*v*) are such that the probability of observing *v ≥ v*′ when there is no outbreak, *p*_01_, is less than the probability of observing *v ≥ v*′ when there is an ongoing outbreak, *p*_11_.

If *v* < *v*′, then we interpret this as meaning that there is no outbreak, while if *v* ≥ *v*′, then we suspect an ongoing outbreak and take appropriate countermeasures. The countermeasures could take many forms. For example, standing water can be drained from containers to reduce the population of mosquito larvae, and large-scale spraying of pesticides can be carried out. These actions directly target the vector population, however, which means that they would necessarily raise the value of *q*_10_ when applied. Since we assume *q*_10_ to be constant in the present treatment, we do not consider these types of interventions for this example. Rather, we consider interventions that lessen the probability of an individual contracting the pathogen, such as advising people to stay indoors and to use insect repellent when they must go outside.

These countermeasures have associated costs and benefits. By remaining indoors due to a public health advisory, people are foregoing activities that they would have otherwise engaged in. The use of insect repellent requires a non-negligible amount of money and time. But by implementing these countermeasures, we reduce the number of human infections due to the outbreak, thereby lessening pathogen-related costs due to morbidity and mortality.

The final step, then, is to understand how the aforementioned costs and benefits manifest in the model parameters *k* and *c*. In this example, *k* measures the costs of the intervention plus the remaining costs of the pathogen despite the intervention. By contrast, *c* measures the costs of the pathogen given that no intervention is applied. If we are able to reconcile all types of costs using a common cost unit, then *K* = *k/c* is a dimensionless number, and Equations 5 tell us if we should intervene, given a particular viral RNA threshold, *v*′. For pathogens such as WNV and EEEV, surveillance is routinely performed, and interventions are helpful for mitigating morbidity and mortality. Therefore, *v*′ must be chosen to be not too low and not too high, so that Equations 5 give us *Y*_0_ = 0 and *Y*_1_ = 1. But what value of *v*′ minimizes *L*_1_(*S*_1_, *K*; *Y*_0_, *Y*_1_)? This value of *v*′, which we denote *v*\*, is the optimal viral RNA threshold for triggering intervention. Figure S3 demonstrates the calculation of *v*\*.

**Figure S3:**
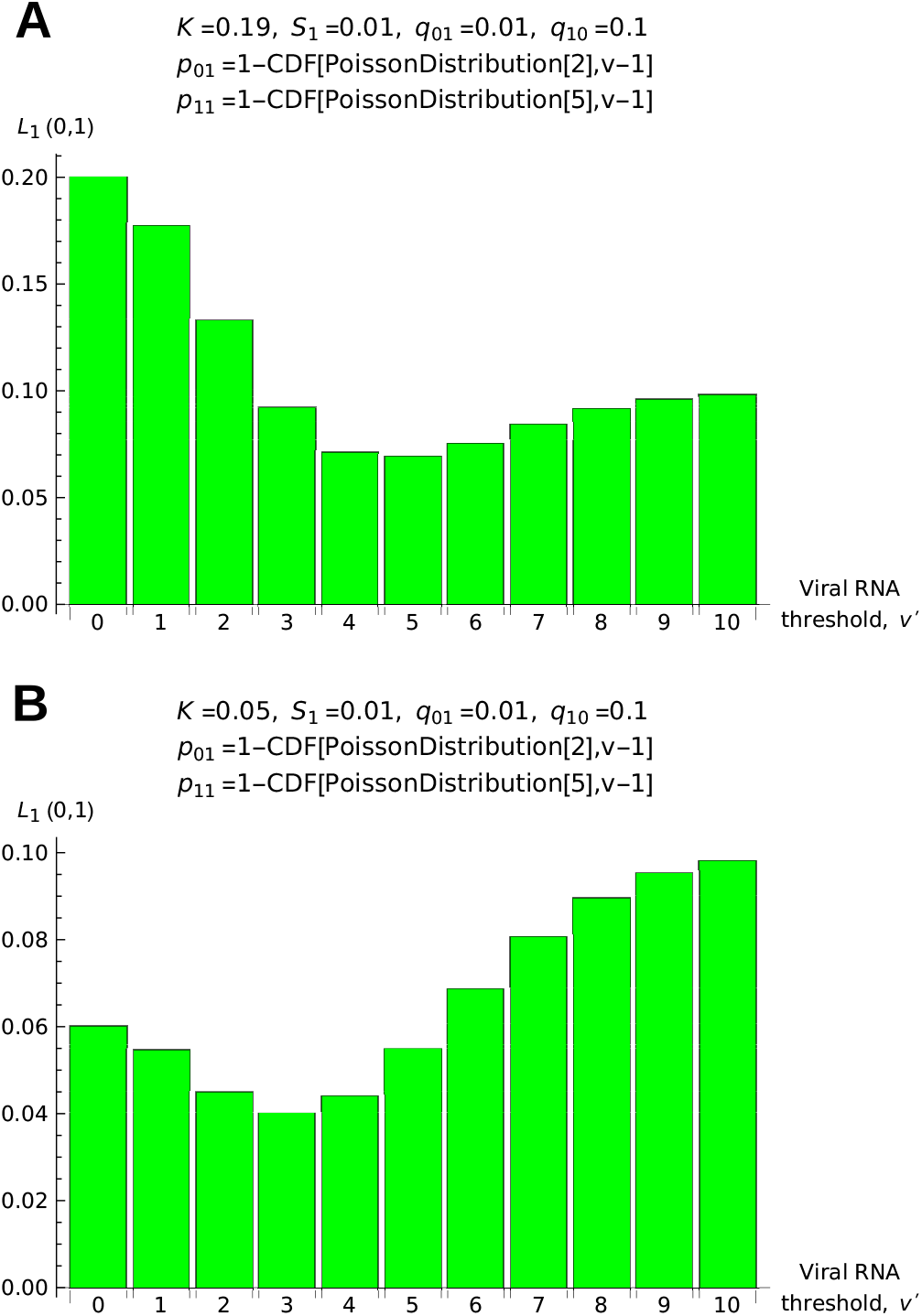
Choosing the optimal viral RNA threshold. When trapping and testing mosquitoes for arboviruses, consider that we are using only the number of infected mosquitoes in the most recent sample to inform whether we should intervene. How many infected mosquitoes, *v*′, in the most recent sample should be required to trigger an alarm? This is determined by choosing the value of *v*′ = *v*\* that minimizes the loss function, *L*_1_(0, 1). Intervention costs are higher in (**A**), where *v*\* = 5, than in (**B**), where *v*\* = 3. In (**A**), we should intervene if and only if there are five or more infected mosquitoes in the most recent sample, while in (**B**), we should intervene if and only if there are three or more infected mosquitoes in the most recent sample. (*K*: normalized intervention cost; *S*_1_: normalized surveillance cost per unit time using one bit of data; *q*_01_: probability of switching from normal to abnormal; *q*_10_: probability of switching from abnormal to normal; *p*_01_: probability of observing 1 while normal; *p*_11_: probability of observing 1 while abnormal.)

### 3 Surveillance using two bits of data

The unnormalized expected cost per unit time given that we use the two most recent bits of data, 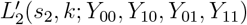, can be expressed as

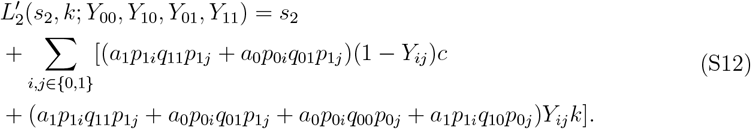

The terms in the summation in Equation (S12) have a simple interpretation. If we do not intervene at time *t* and the system is in the abnormal state at time *t*, then we incur a delayed cost, *c*. Considering the two most recent bits of data, there are eight ways that this can happen:

- The system can be in the abnormal state at time *t* − 1, be measured as bit *i* at time *t* − 1, remain in the abnormal state at time *t*, be measured as bit *j* at time *t*, and not be acted on at time *t*. This sequence of events occurs with probability *a*_1_*p*_1*i*_*q*_11_*p*_1*j*_(1 *−Y*_*ij*_), and *i* and *j* can each be either 0 or 1.
- The system can be in the normal state at time *t* −1, be measured as bit *i* at time *t* −1, transition to the abnormal state at time *t*, be measured as bit *j* at time *t*, and not be acted on at time *t*. This sequence of events occurs with probability *a*_0_*p*_0*i*_*q*_01_*p*_1*j*_(1 *−Y*_*ij*_), and *i* and *j* can each be either 0 or 1.

If we intervene at time *t*, then we incur an immediate cost, *k*. Considering the two most recent bits of data, there are sixteen ways that this can happen:

- The system can be in the abnormal state at time *t* − 1, be measured as bit *i* at time *t* − 1, remain in the abnormal state at time *t*, be measured as bit *j* at time *t*, and be acted on at time *t*. This sequence of events occurs with probability *a*_1_*p*_1*i*_*q*_11_*p*_1*j*_*Y*_*ij*_, and *i* and *j* can each be either 0 or 1.
- The system can be in the normal state at time *t* − 1, be measured as bit *i* at time *t* − 1, transition to the abnormal state at time *t*, be measured as bit *j* at time *t*, and be acted on at time *t*. This sequence of events occurs with probability *a*_0_*p*_0*i*_*q*_01_*p*_1*j*_*Y*_*ij*_, and *i* and *j* can each be either 0 or 1.
- The system can be in the normal state at time *t* −1, be measured as bit *i* at time *t* −1, remain in the normal state at time *t*, be measured as bit *j* at time *t*, and be acted on at time *t*. This sequence of events occurs with probability *a*_0_*p*_0*i*_*q*_00_*p*_0*j*_*Y*_*ij*_, and *i* and *j* can each be either 0 or 1.
- The system can be in the abnormal state at time *t* − 1, be measured as bit *i* at time *t* − 1, transition to the normal state at time *t*, be measured as bit *j* at time *t*, and be acted on at time *t*. This sequence of events occurs with probability *a*_1_*p*_1*i*_*q*_10_*p*_0*j*_*Y*_*ij*_, and *i* and *j* can each be either 0 or 1.

Letting *S*_2_ = *s*_2_*/c* and 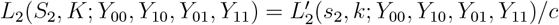 in Equation (S12), we obtain the normalized expected cost per unit time:

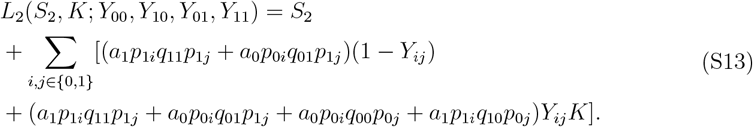

As shorthand notation, we define

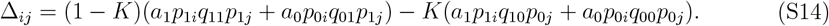

Rearranging Equation (S13), and using Equation (S14), we obtain

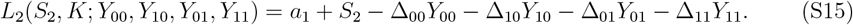

The task at hand is to use Equation (S15) and the observed bit sequence to guide our intervention strategy.

For some applications, surveillance using the two most recent bits of data can be detrimental. In Figure S4, for a particular sequence of system states and observed bits, we compare the strategy to intervene if and only if we observe 1 to the strategy to intervene if and only if we observe 11. For the latter strategy, the number of Type I errors is reduced but the number of Type II errors is substantially increased, such that requiring an observation of 11 in succession for deciding to intervene is counterproductive. In Figure S5, for the same sequence of system states and observed bits, we compare the strategy to intervene if and only if we observe 1 to the strategy to not intervene if and only if we observe 00. For the latter strategy, the number of Type II errors is reduced but the number of Type I errors is substantially increased, such that requiring an observation of 00 in succession for deciding to not intervene is counterproductive.

Another example shows how surveillance using the two most recent bits of data can be optimal (Table 2). If intervention costs are low, then it might be beneficial to not intervene if and only if both of the most recent bits are 0. By observing 00 in succession, we are more confident that the system is in the normal state and that intervention is useless. If intervention costs are high, then it might be beneficial to intervene if and only if both of the most recent bits are 1. By observing 11 in succession, we are more confident that the system is in the abnormal state and that intervention is worthwhile.

**Figure S4:**
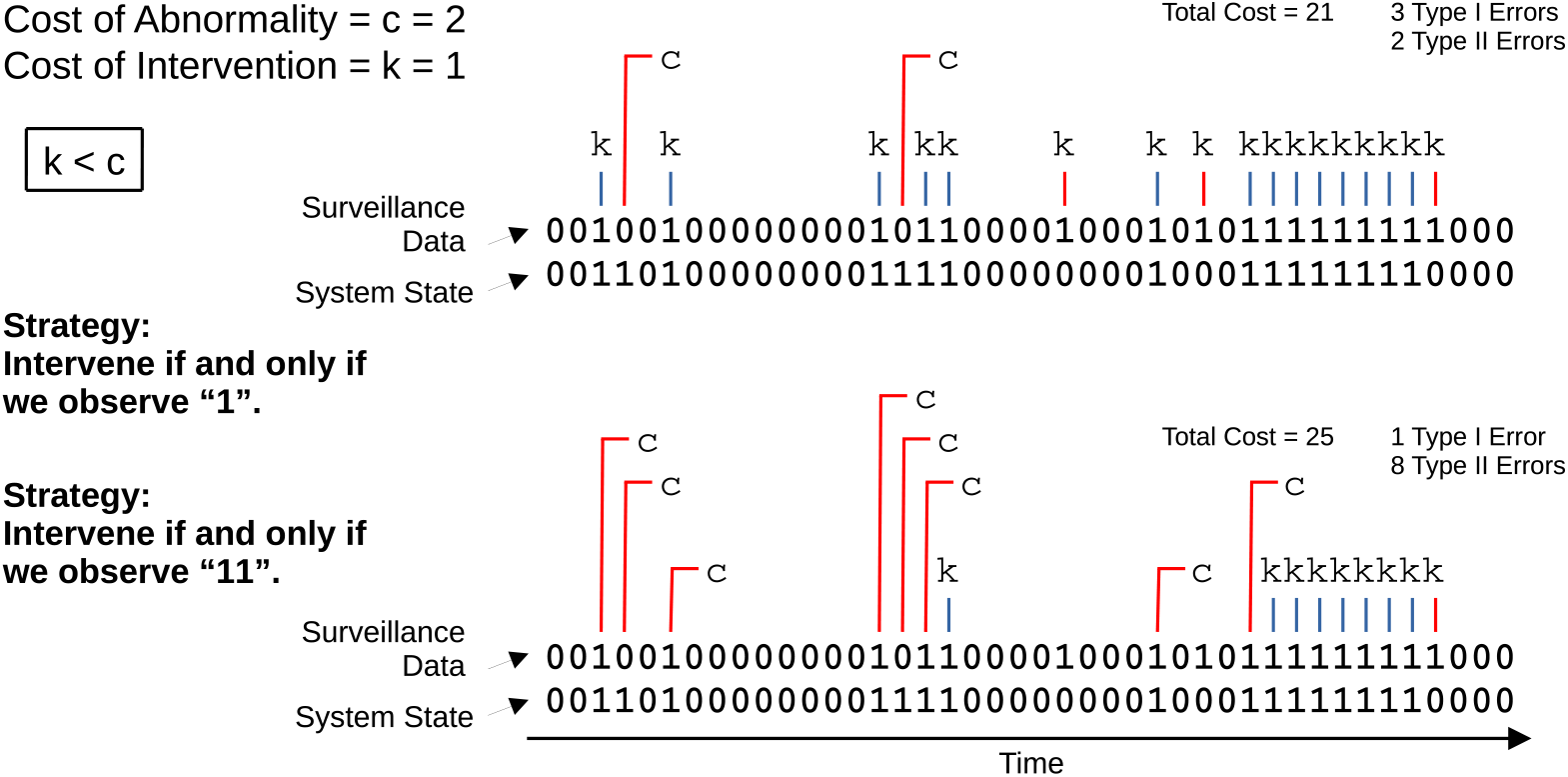
Requiring reinforcement for intervening. For this particular sequence of system state values and measurements, we compare the strategy to intervene if and only if we observe 1 versus the strategy to intervene if and only if we observe 11 in succession. For the latter strategy, there are two fewer Type I errors but six more Type II errors. The additional Type II errors mean that the latter strategy would have performed worse than the former for this realization of the dynamics.

**Figure S5:**
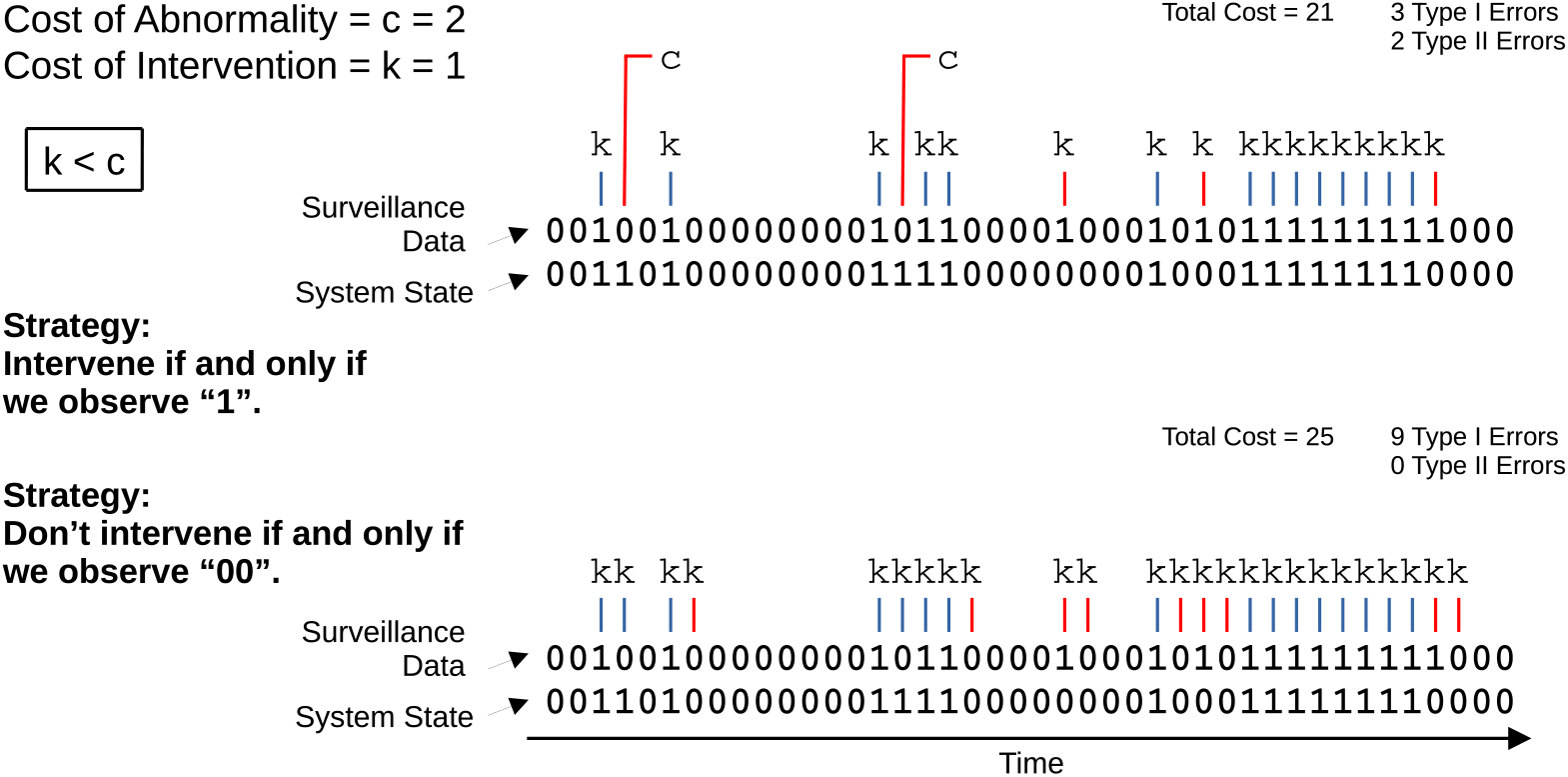
Requiring reinforcement for not intervening. For this particular sequence of system state values and measurements, we compare the strategy to intervene if and only if we observe 1 versus the strategy to not intervene if and only if we observe 00 in succession. For the latter strategy, there are two fewer Type II errors but six more Type I errors. The additional Type I errors mean that the latter strategy would have performed worse than the former for this realization of the dynamics.

**Table 2:**
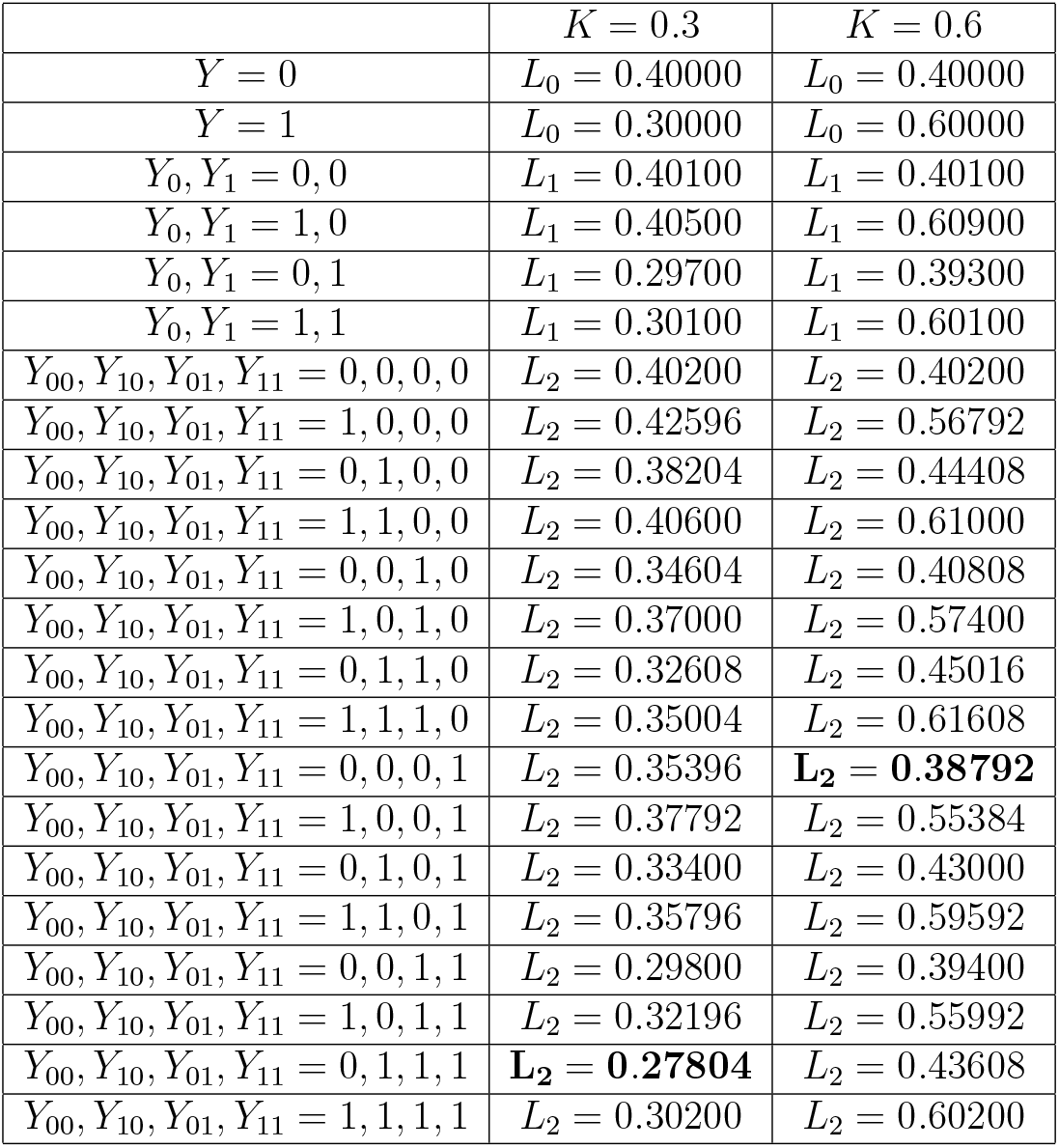
Example of optimizing surveillance and intervention using the two most recent bits of data. We set *q*_01_ = 0.2, *q*_10_ = 0.3, *p*_01_ = 0.2, *p*_11_ = 0.5, *S*_1_ = 0.001, and *S*_2_ = 0.002.

| | $K = 0.3$ | $K = 0.6$ |
| --- | --- | --- |
| $Y = 0$ | $L_0 = 0.40000$ | $L_0 = 0.40000$ |
| $Y = 1$ | $L_0 = 0.30000$ | $L_0 = 0.60000$ |
| $Y_0, Y_1 = 0, 0$ | $L_1 = 0.40100$ | $L_1 = 0.40100$ |
| $Y_0, Y_1 = 1, 0$ | $L_1 = 0.40500$ | $L_1 = 0.60900$ |
| $Y_0, Y_1 = 0, 1$ | $L_1 = 0.29700$ | $L_1 = 0.39300$ |
| $Y_0, Y_1 = 1, 1$ | $L_1 = 0.30100$ | $L_1 = 0.60100$ |
| $Y_{00}, Y_{10}, Y_{01}, Y_{11} = 0, 0, 0, 0$ | $L_2 = 0.40200$ | $L_2 = 0.40200$ |
| $Y_{00}, Y_{10}, Y_{01}, Y_{11} = 1, 0, 0, 0$ | $L_2 = 0.42596$ | $L_2 = 0.56792$ |
| $Y_{00}, Y_{10}, Y_{01}, Y_{11} = 0, 1, 0, 0$ | $L_2 = 0.38204$ | $L_2 = 0.44408$ |
| $Y_{00}, Y_{10}, Y_{01}, Y_{11} = 1, 1, 0, 0$ | $L_2 = 0.40600$ | $L_2 = 0.61000$ |
| $Y_{00}, Y_{10}, Y_{01}, Y_{11} = 0, 0, 1, 0$ | $L_2 = 0.34604$ | $L_2 = 0.40808$ |
| $Y_{00}, Y_{10}, Y_{01}, Y_{11} = 1, 0, 1, 0$ | $L_2 = 0.37000$ | $L_2 = 0.57400$ |
| $Y_{00}, Y_{10}, Y_{01}, Y_{11} = 0, 1, 1, 0$ | $L_2 = 0.32608$ | $L_2 = 0.45016$ |
| $Y_{00}, Y_{10}, Y_{01}, Y_{11} = 1, 1, 1, 0$ | $L_2 = 0.35004$ | $L_2 = 0.61608$ |
| $Y_{00}, Y_{10}, Y_{01}, Y_{11} = 0, 0, 0, 1$ | $L_2 = 0.35396$ | **$L_2 = 0.38792$** |
| $Y_{00}, Y_{10}, Y_{01}, Y_{11} = 1, 0, 0, 1$ | $L_2 = 0.37792$ | $L_2 = 0.55384$ |
| $Y_{00}, Y_{10}, Y_{01}, Y_{11} = 0, 1, 0, 1$ | $L_2 = 0.33400$ | $L_2 = 0.43000$ |
| $Y_{00}, Y_{10}, Y_{01}, Y_{11} = 1, 1, 0, 1$ | $L_2 = 0.35796$ | $L_2 = 0.59592$ |
| $Y_{00}, Y_{10}, Y_{01}, Y_{11} = 0, 0, 1, 1$ | $L_2 = 0.29800$ | $L_2 = 0.39400$ |
| $Y_{00}, Y_{10}, Y_{01}, Y_{11} = 1, 0, 1, 1$ | $L_2 = 0.32196$ | $L_2 = 0.55992$ |
| $Y_{00}, Y_{10}, Y_{01}, Y_{11} = 0, 1, 1, 1$ | **$L_2 = 0.27804$** | $L_2 = 0.43608$ |
| $Y_{00}, Y_{10}, Y_{01}, Y_{11} = 1, 1, 1, 1$ | $L_2 = 0.30200$ | $L_2 = 0.60200$ |

We have already worked out the conditions for when surveillance using one bit is superior to no surveillance. When we include the possibility of using two bits of data to guide our intervention, determination of the optimal surveillance and intervention strategy becomes more intricate. We must still consider how surveillance using one bit performs versus no surveillance, as shown in Figure 3 and Figure 4A. We must similarly determine if surveillance using two bits is superior to no surveillance, and we must determine if surveillance using two bits is superior to surveillance using one bit.

When comparing surveillance using two bits to no surveillance, there are multiple comparisons that must be done separately. First, we compare the strategy to intervene if and only if we observe 11 to never intervening. If *K* is larger than a certain value, which we denote *K*_0+_, then we should never intervene. *K*_0+_ is given by solving *L*_2_(0, *K*_0+_; 0, 0, 0, 1) = *L*_0_(*K*_0+_; 0). We find

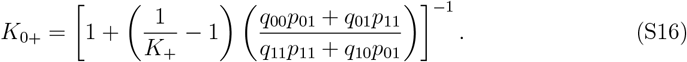

If *K* > *K*_0+_, then the expected cost from reacting to false positives whenever we observe 11 exceeds any benefit from averting the threat.

Similarly, we compare the strategy to intervene if and only if we observe 11 to always intervening. If *K* is smaller than a certain value, which we denote *K*_0−_, then we should always intervene. *K*_0−_ is given by solving *L*_2_(0, *K*_0−_; 0, 0, 0, 1) = *L*_0_(*K*_0−_; 1). We find

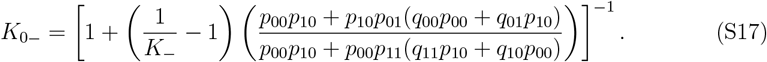

If *K* < *K*_0−_, then the expected cost from dismissing false negatives whenever we do not observe 11 exceeds any benefit from not intervening when there is no threat.

If *K*_0−_ < *K* < *K*_0+_, then using two bits and intervening if and only if we observe 11 might be justified if surveillance costs are sufficiently low. If we use surveillance, then we incur a cost per unit time equal to *L*_2_(*S*_2_, *K*; 0, 0, 0, 1). First, consider that *K* > *a*_1_. If we don’t use surveillance, then the optimal strategy is to never intervene, and we incur a cost per unit time equal to *L*_0_(*K*; 0). We set *L*_0_(*K*; 0) = *L*_2_(*S*_0+_, *K*; 0, 0, 0, 1) and solve for *S*_0+_:

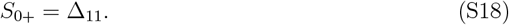

If *S*_2_ < *S*_0+_, then the benefit of appropriately intervening whenever we observe 11 outweighs the cost of surveillance, and surveillance using two bits is beneficial relative to never intervening. If *S*_2_ > *S*_0+_, then surveillance using two bits is too expensive relative to never intervening. From Equation (S18), *S*_0+_ is plotted versus *K* as the upper right boundary of the triangular region in Figure 4B.

Next, consider that *K* < *a*_1_. If we don’t use surveillance, then the optimal strategy is to always intervene, and we incur a cost per unit time equal to *L*_0_(*K*; 1). We set *L*_0_(*K*; 1) = *L*_2_(*S*_0−_, *K*; 0, 0, 0, 1) and solve for *S*_0−_:

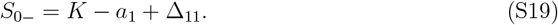

If *S*_2_ < *S*_0−_, then the benefit of appropriately intervening whenever we observe 11 outweighs the cost of surveillance, and surveillance using two bits is beneficial relative to always intervening. If *S*_2_ > *S*_0−_, then surveillance using two bits is too expensive relative to always intervening. From Equation (S19), *S*_0−_ is plotted versus *K* as the upper left boundary of the triangular region in Figure 4B.

Next, we compare the strategy to not intervene if and only if we observe 00 to never intervening. If *K* is larger than a certain value, which we denote *K*_1+_, then we should never intervene. *K*_1+_ is given by solving *L*_2_(0, *K*_1+_; 0, 1, 1, 1) = *L*_0_(*K*_1+_; 0). We find

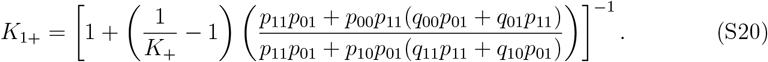

If *K* > *K*_1+_, then the expected cost from reacting to false positives whenever we do not observe 00 exceeds any benefit from averting the threat.

Similarly, we compare the strategy to not intervene if and only if we observe 00 to always intervening. If *K* is smaller than a certain value, which we denote *K*_1−_, then we should always intervene. *K*_1−_ is given by solving *L*_2_(0, *K*_1−_; 0, 1, 1, 1) = *L*_0_(*K*_1−_; 1). We find

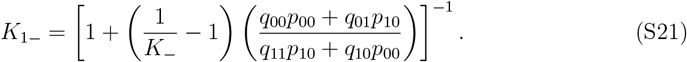

If *K* < *K*_1−_, then the expected cost from dismissing false negatives whenever we observe 00 exceeds any benefit from not intervening when there is no threat.

If *K*_1−_ < *K* < *K*_1+_, then using two bits and not intervening if and only if we observe 00 might be justified if surveillance costs are sufficiently low. If we use surveillance, then we incur a cost per unit time equal to *L*_2_(*S*_2_, *K*; 0, 1, 1, 1). First, consider that *K* > *a*_1_. If we don’t use surveillance, then the optimal strategy is to never intervene, and we incur a cost per unit time equal to *L*_0_(*K*; 0). We set *L*_0_(*K*; 0) = *L*_2_(*S*_1+_, *K*; 0, 1, 1, 1) and solve for *S*_1+_:

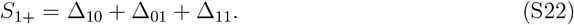

If *S*_2_ < *S*_1+_, then the benefit of appropriately intervening whenever we do not observe 00 outweighs the cost of surveillance, and surveillance using two bits is beneficial relative to always intervening. If *S*_2_ *> S*_1+_, then surveillance using two bits is too expensive relative to always intervening. From Equation (S22), *S*_1+_ is plotted versus *K* as the upper right boundary of the triangular region in Figure 4C.

Next, consider that *K* < *a*_1_. If we don’t use surveillance, then the optimal strategy is to always intervene, and we incur a cost per unit time equal to *L*_0_(*K*; 1). We set *L*_0_(*K*; 1) = *L*_2_(*S*_1−_, *K*; 0, 1, 1, 1) and solve for *S*_1−_:

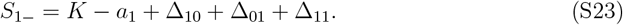

If *S*_2_ < *S*_1−_, then the benefit of appropriately intervening whenever we do not observe 00 outweighs the cost of surveillance, and surveillance using two bits is beneficial relative to always intervening. If *S*_2_ *> S*_1−_, then surveillance using two bits is too expensive relative to always intervening. From Equation (S23), *S*_1−_ is plotted versus *K* as the upper left boundary of the triangular region in Figure 4C.

We must also determine if surveillance using two bits is superior to surveillance using one bit. If *K* is between two values, which we denote *K*_2+_ and *K*_2−_, then we should intervene if and only if we observe 1. *K*_2+_ is equal to the value of *K* for which *L*_2_(*S*_1_, *K*_2+_; 0, 0, 0, 1) = *L*_1_(*S*_1_, *K*_2+_; 0, 1). We find

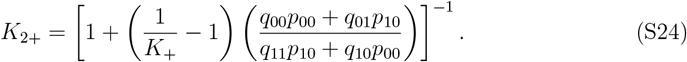

*K*_2−_ is equal to the value of *K* for which *L*_2_(*S*_1_, *K*_2−_; 0, 1, 1, 1) = *L*_1_(*S*_1_, *K*_2−_; 0, 1). We find

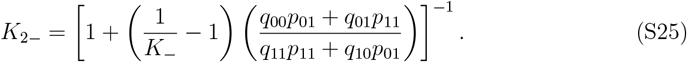

For *K* > *K*_2+_, surveillance using two bits is superior to surveillance using one bit if *S*_2_ *− S*_1_ < *S*_2+_, where *S*_2+_ is given by *L*_2_(*S*_2+_, *K*; 0, 0, 0, 1) = *L*_1_(0, *K*; 0, 1). We have

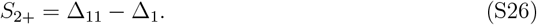

For *K* < *K*_2−_, surveillance using two bits is superior to surveillance using one bit if *S*_2_ *− S*_1_ *< S*_2−_, where *S*_2−_ is given by *L*_2_(*S*_2−_, *K*; 0, 1, 1, 1) = *L*_1_(0, *K*; 0, 1). We have

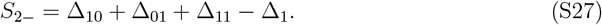

From Equations (S26) and (S27), *S*_2+_ and *S*_2−_ are plotted versus *K* as the right and left boundary lines, respectively, in Figure 4D.

The understanding of surveillance using two bits is summarized concisely in the four panels of Figure 4. What can we learn from this plot? For simplicity in the descriptions that follow, suppose that all surveillance costs are negligible.

One might first ask: If we must decide between no surveillance and surveillance using two bits, where we intervene if and only if we observe 11, then what should we do? There are two *K* values, *K*_0+_ and *K*_0−_, that answer this question. For context, it is insightful to compare these quantities to *K*_+_ and *K*_−_, respectively. From inspecting Equation (S16), *K*_0+_ is necessarily larger than *K*_+_. This is because when we require observing 11 to intervene, we are reducing the rate of false positives, so for interventions for which *K*_+_ < *K* < *K*_0+_, false positives are easier to tolerate, and it makes sense to have surveillance. From inspecting Equation (S17), *K*_0−_ is necessarily larger than *K*_−_. This is because when we require observing 11 to intervene, we are increasing the rate of false negatives, so for interventions for which *K*_−_ < *K* < *K*_0−_, it makes sense to always intervene, thereby averting any costs due to missed detection.

One might then ask: If we must decide between no surveillance and surveillance using two bits, where we do not intervene if and only if we observe 00, then what should we do? There are two *K* values, *K*_1+_ and *K*_1−_, that answer this question. From inspecting Equation (S20), *K*_1+_ is necessarily smaller than *K*_+_. This is because when we require observing 00 to not intervene, we are increasing the rate of false positives, so for interventions for which *K*_1+_ < *K* < *K*_+_, it makes sense to never intervene, thereby averting any costs due to unwarranted intervention. From inspecting Equation (S21), *K*_1−_ is necessarily smaller than *K*_−_. This is because when we require observing 00 to not intervene, we are reducing the rate of false negatives, so for interventions for which *K*_1−_ < *K* < *K*_−_, false negatives are easier to tolerate, and it makes sense to have surveillance.

One might also ask: If we must decide between surveillance using one bit and surveillance using two bits, then what should we do? There are two *K* values, *K*_2+_ and *K*_2−_, that answer this question. From inspecting Equations (S24) and (S25), *K*_2+_ is necessarily smaller than *K*_+_, and *K*_2−_ is necessarily larger than *K*_−_. Therefore, if *K*_+_ < *K* < 1, then we should never intervene, and if 0 < *K* < *K*_−_, then we should always intervene. Further, if *K*_2+_ < *K* < *K*_+_, then intervention is sufficiently costly that surveillance using two bits—with the strategy to intervene if and only if we observe 11—is helpful for reducing false positives and unnecessary intervention costs. Also, if *K*_−_ < *K* < *K*_2−_, then intervention is sufficiently cheap that surveillance using two bits—with the strategy to not intervene if and only if we observe 00—is helpful for reducing false negatives and costs due to missed detection. But if *K*_2−_ < *K* < *K*_2+_, then surveillance using two bits is inferior to surveillance using one bit. For these values of *K*, when considering the strategy to intervene if and only if we observe 11, the benefit of averting false positives is outweighed by the cost of false negatives. Also for these values of *K*, when considering the strategy to not intervene if and only if we observe 00, the benefit of averting false negatives is outweighed by the cost of false positives.

#### 3.1 Example using two bits of data

Using two measurements in succession, how should we decide whether or not to intervene? It is worth noting that a proper decision rule in this setting would use all available information, which would include the viral RNA abundance measured in the samples from both the previous week and the current week. But for applying this model, we use only a single bit of data to represent each weekly measurement. If *v* ≥ *v*′ for the previous week, then the previous week’s observation is 1, and 0 otherwise. If *v* ≥ *v*′ for the current week, then the current week’s observation is 1, and 0 otherwise. Given a particular viral RNA threshold, *v*′, Equations 8 determine the ideal intervention strategy, and we must choose the optimal value of *v*′, given by *v*\*, such that *L*_2_(*S*_2_, *K*; *Y*_00_, *Y*_10_, *Y*_01_, *Y*_11_) is minimized. A simple demonstration considering strategies that use either one or two bits of data, again in the context of surveillance for arboviruses, is shown in Figure S6.

### 4 Example using three bits of data

When considering surveillance using more than two bits of data, calculation of the optimal strategy is more complicated. For example, consider two possible bit sequences: 001 and 110. In the former, the most recent measurement is 1, while the two previous measurements are both 0. In the latter, the most recent measurement is 0, while the two previous measurements are both 1. For determining the intervention strategy, does the most recent observation of 1 or 0 outweigh the two previous observations of 00 or 11, or vice versa?

Figure S7 shows two possibilities for different parameter sets. First, consider Figure S7A for the case of large intervention costs (inside the gray region). A 1 in the most recent measurement outweighs 0 readings in both of the two previous measurements, and we should intervene if we observe 001. Also, a 0 in the most recent measurement outweighs two 1 readings in both of the two previous measurements, and we should not intervene if we observe 110. Thus, the analysis simplifies: Since a measurement of 001 warrants intervention, measurements of 011, 101, or 111 also warrant intervention. Furthermore, since a measurement of 110 should not induce an intervention, measurements of 100, 010, or 000 also should not induce an intervention. The conclusion is that the optimal strategy is to use only the most recent bit of data, and we should intervene if and only if this bit is 1.

**Figure S6:**
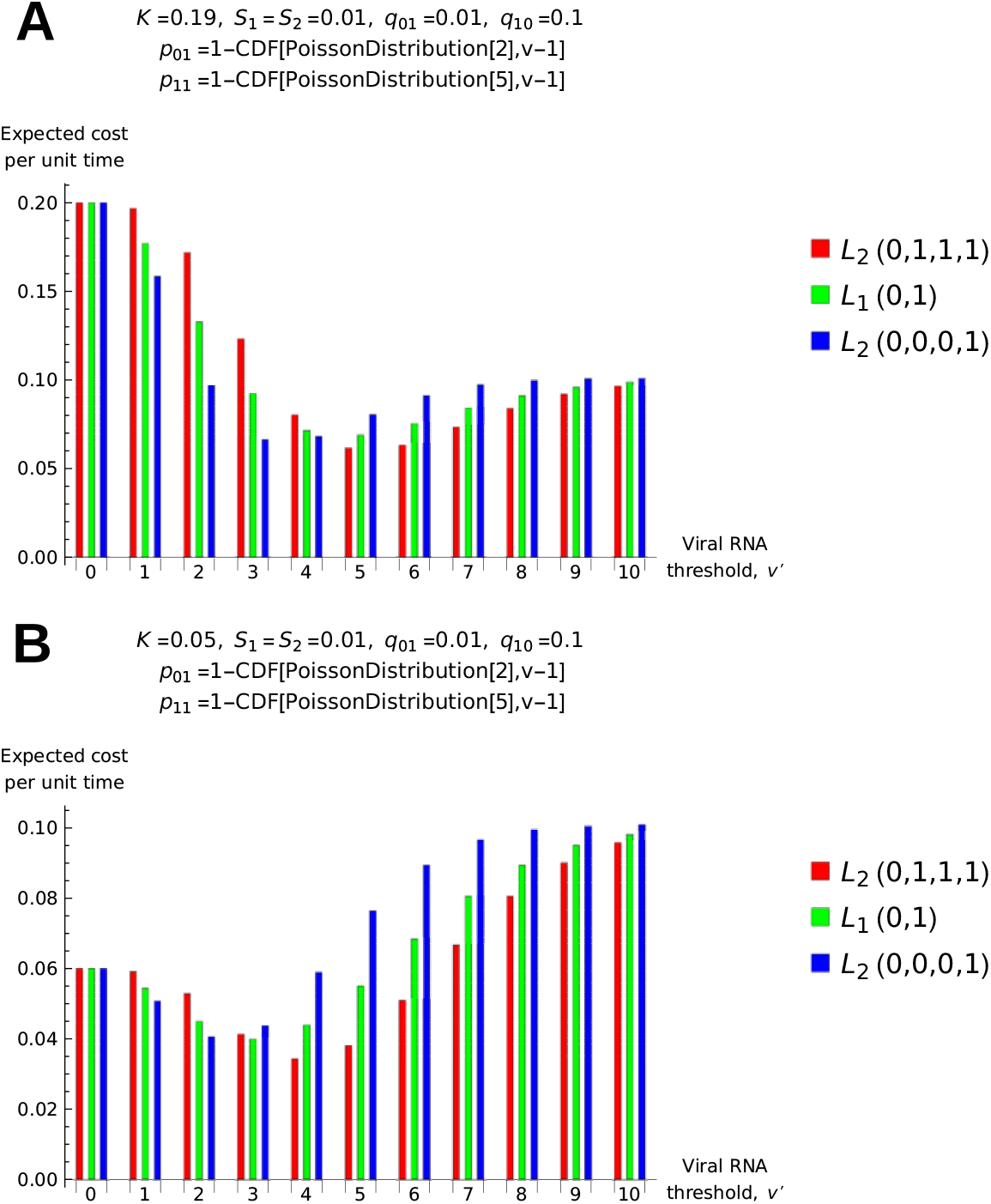
Choosing the optimal viral RNA threshold and the optimal number of bits to use. The parameter values are the same as in Figure S3. However, here, we consider two additional strategies: intervene if and only if the two most recent bits are 1, and intervene if and only if at least one of the two most recent bits is 1. The corresponding loss functions are *L*_2_(0, 0, 0, 1) and *L*_2_(0, 1, 1, 1), respectively. In both (**A**) and (**B**), the strategy to use only a single bit of data is outperformed by the strategy to intervene if and only if at least one of the two most recent bits is 1. In (**A**), we have *v*\* = 5, and we should intervene if and only if at least one of the two most recent samples contained five or more mosquitoes. In (**B**), we have *v*\* = 4, and we should intervene if and only if at least one of the two most recent samples contained four or more mosquitoes. (*K*: normalized intervention cost; *S*_1_ = *S*_2_: normalized surveillance cost per unit time using one or two bits of data; *q*_01_: probability of switching from normal to abnormal; *q*_10_: probability of switching from abnormal to normal; *p*_01_: probability of observing 1 while normal; *p*_11_: probability of observing 1 while abnormal.)

**Figure S7:**
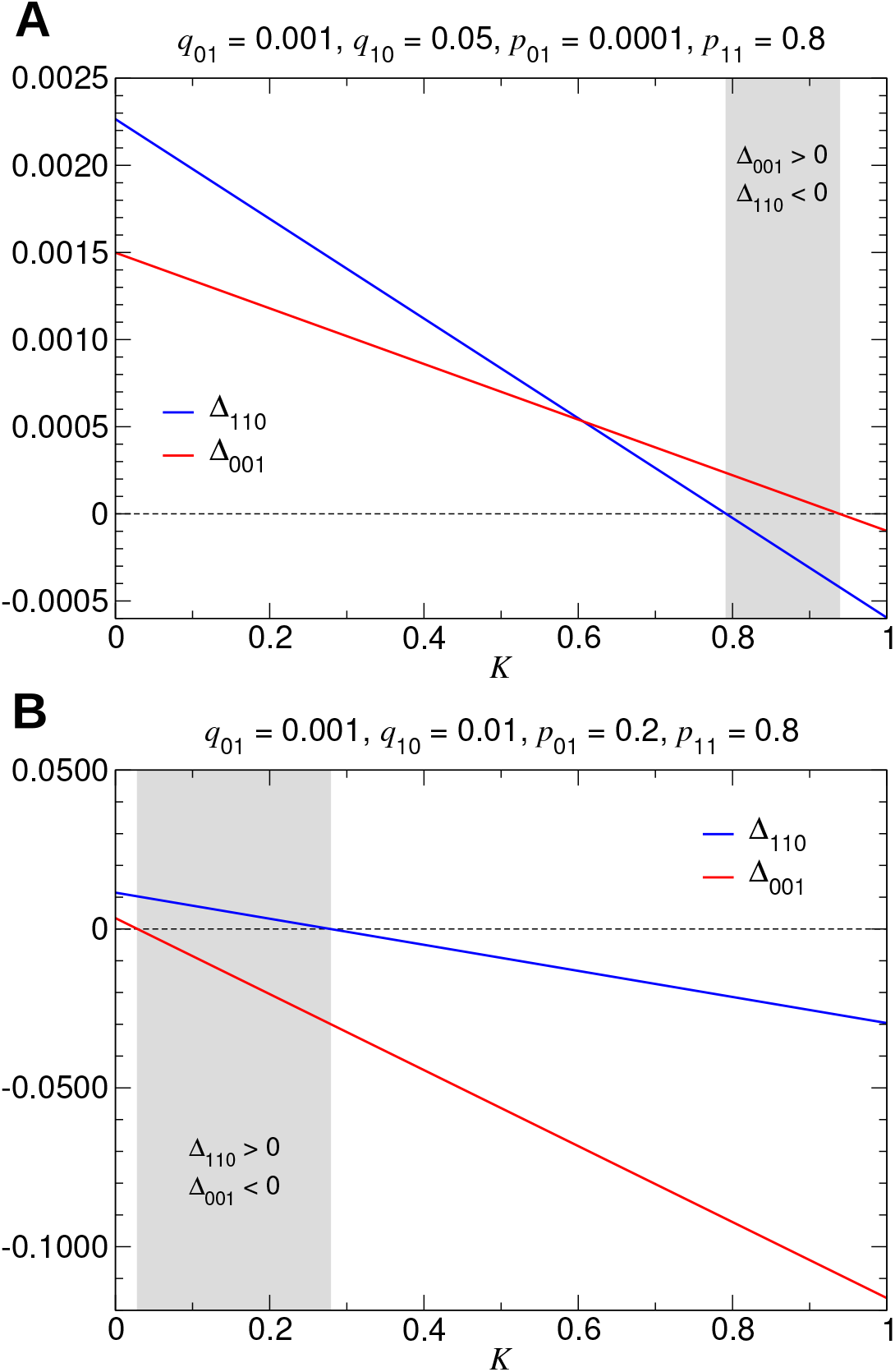
Surveillance using three bits of data. In (**A**), for intervention costs within the gray region, we should intervene if we observe 001 and not intervene if we observe 110. In (**B**), for intervention costs within the gray region, we should intervene if we observe 110 and not intervene if we observe 001. This example illustrates that optimization of surveillance and intervention using more than two bits of data can be intricate. (*q*_01_: probability of switching from normal to abnormal; *q*_10_: probability of switching from abnormal to normal; *p*_01_: probability of observing 1 while normal; *p*_11_: probability of observing 1 while abnormal.)

**Figure S8:**
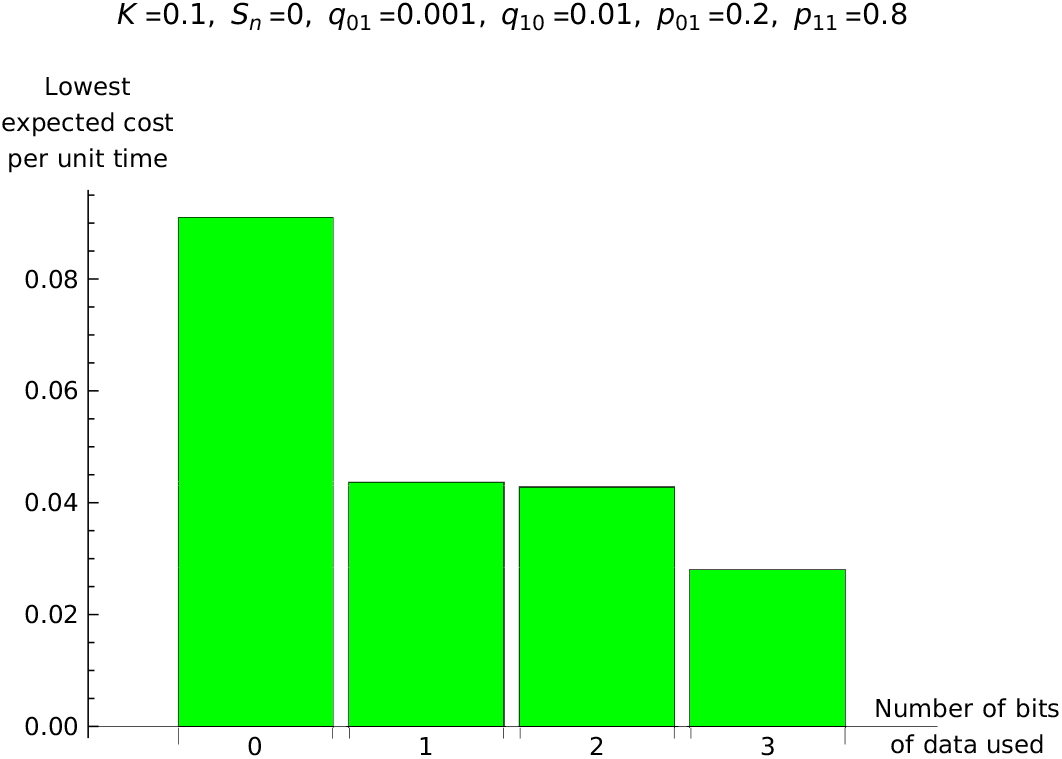
Determining the optimal amount of data to use. Consider the lowest expected cost per unit time that is achievable by using zero, one, two, or three bits of data in the decision to intervene. Using one bit of data yields a large improvement over no surveillance, using two bits of data yields a marginal improvement over using one bit of data, and using three bits of data yields a large improvement over using two bits of data. (*K*: normalized intervention cost; *S*_*n*_: normalized surveillance cost per unit time using *n* bits of data; *q*_01_: probability of switching from normal to abnormal; *q*_10_: probability of switching from abnormal to normal; *p*_01_: probability of observing 1 while normal; *p*_11_: probability of observing 1 while abnormal.)

By contrast, consider Figure S7B for small intervention costs (inside the gray region). Two previous 1 readings in succession outweigh a 0 in the most recent measurement, and we should intervene if we observe 110. Also, two previous 0 readings in succession outweigh a 1 in the most recent measurement, and we should not intervene if we observe 001. Using the three most recent bits of data, the optimal strategy is that we should intervene if and only if we observe 110, 101, 011, or 111. This example shows that care must be taken when determining the optimal strategy if more than two bits of data are being used.

A further consideration is the number of bits of data to use for deciding whether to intervene. For the same parameter values as in Figure S7B, Figure S8 shows the lowest cost per unit time that is achievable when using zero, one, two, or three bits of data. Using zero bits of data corresponds to no surveillance and no intervention, and the resulting expected cost per unit time provides a benchmark. By using just a single bit of data, and by intervening if and only if we observe 1, we achieve a > 50% reduction in the expected cost per unit time.

Using two bits of data and intervening if and only if we observe 11 delivers only a marginal benefit over using one bit of data. However, using three bits of data delivers a dramatic improvement in performance over using two bits of data. Determination of the optimal number of bits of data to use is therefore not always an intuitive exercise, and care must be taken in doing so.

